# Subcutaneous REGEN-COV Antibody Combination for Covid-19 Prevention

**DOI:** 10.1101/2021.06.14.21258567

**Authors:** Meagan P. O’Brien, Eduardo Forleo-Neto, Bret J. Musser, Flonza Isa, Kuo-Chen Chan, Neena Sarkar, Katharine J. Bar, Ruanne V. Barnabas, Dan H. Barouch, Myron S. Cohen, Christopher B. Hurt, Dale R. Burwen, Mary A. Marovich, Peijie Hou, Ingeborg Heirman, John D. Davis, Kenneth C. Turner, Divya Ramesh, Adnan Mahmood, Andrea T. Hooper, Jennifer D. Hamilton, Yunji Kim, Lisa A. Purcell, Alina Baum, Christos A. Kyratsous, James Krainson, Richard Perez-Perez, Rizwana Mohseni, Bari Kowal, A. Thomas DiCioccio, Neil Stahl, Leah Lipsich, Ned Braunstein, Gary Herman, George D. Yancopoulos, David M. Weinreich, for the Covid-19 Phase 3 Prevention Trial Team

**Affiliations:** Regeneron Pharmaceuticals, Inc., Tarrytown, NY, USA; Department of Medicine, University of Pennsylvania, Philadelphia, PA, USA; Department of Microbiology, University of Pennsylvania, Philadelphia, PA, USA; Department of Global Health, University of Washington, Seattle, WA, USA; Division of Allergy and Infectious Diseases, University of Washington, Seattle, WA, USA; Department of Epidemiology, University of Washington, Seattle, WA, USA; Vaccine and Infectious Diseases Division, Fred Hutchinson Cancer Research Center, Seattle, WA, USA; Center for Virology and Vaccine Research, Beth Israel Deaconess Medical Center, Harvard Medical School, Boston, MA, USA; Institute for Global Health and Infectious Diseases, University of North Carolina at Chapel Hill, Chapel Hill, NC, USA; National Institute of Allergy and Infectious Diseases, National Institutes of Health, Rockville, MD, USA; Clinical Trials of Florida, LLC, Miami, FL, USA; Medical Research of Westchester, Miami, FL, USA; Catalina Research Institute, LLC, Montclair, CA, USA

## Abstract

**Background:** Casirivimab and imdevimab (REGEN-COV™) markedly reduces risk of hospitalization or death in high-risk individuals with Covid-19. Here we explore the possibility that subcutaneous REGEN-COV prevents SARS-CoV-2 infection and subsequent Covid-19 in individuals at high risk of contracting SARS-CoV-2 by close exposure in a household with a documented SARS-CoV-2–infected individual.

**Methods:** Individuals ≥12 years were enrolled within 96 hours of a household contact being diagnosed with SARS-CoV-2 and randomized 1:1 to receive 1200 mg REGEN-COV or placebo via subcutaneous injection. The primary efficacy endpoint was the proportion of participants without evidence of infection (SARS-CoV-2 RT-qPCR– negative) or prior immunity (seronegative) who subsequently developed symptomatic SARS-CoV-2 infection during a 28-day efficacy assessment period.

**Results:** Subcutaneous REGEN-COV significantly prevented symptomatic SARS-CoV-2 infection compared with placebo (81.4% risk reduction; 11/753 [1.5%] vs. 59/752 [7.8%], respectively; P<0.0001), with 92.6% risk reduction after the first week (2/753 [0.3%] vs. 27/752 [3.6%], respectively). REGEN-COV also prevented overall infections, either symptomatic or asymptomatic (66.4% risk reduction). Among infected participants, the median time to resolution of symptoms was 2 weeks shorter with REGEN-COV vs. placebo (1.2 vs. 3.2 weeks, respectively), and the duration of time with high viral load (>10^4^ copies/mL) was lower (0.4 vs. 1.3 weeks, respectively). REGEN-COV was generally well tolerated.

**Conclusions:** Administration of subcutaneous REGEN-COV prevented symptomatic Covid-19 and asymptomatic SARS-CoV-2 infection in uninfected household contacts of infected individuals. Among individuals who became infected, REGEN-COV reduced the duration of symptomatic disease, decreased maximal viral load, and reduced the duration of detectable virus.

(ClinicalTrials.gov number, NCT04452318.)

## INTRODUCTION

Coronavirus disease 2019 (Covid-19), caused by severe acute respiratory syndrome coronavirus 2 (SARS-CoV-2), first emerged in December 2019 and was declared a global pandemic in March 2020.^1-3^ Casirivimab and imdevimab (REGEN-COV^™^), a monoclonal antibody combination consisting of two neutralizing monoclonal antibodies (administered together) that bind non-competing epitopes of the SARS-CoV-2 spike protein receptor binding domain, retains neutralization potency against circulating SARS-CoV-2 variants of concern in vitro and in vivo (including B.1.1.7, B.1.429, B.1.351, and P.1) and may protect against the selection of resistant variants.^4-6^ In outpatients with Covid-19, REGEN-COV reduced hospitalization or all-cause death by approximately 70%, while rapidly reducing viral load and shortening symptom duration.^7,8^

This study evaluated whether subcutaneously administered REGEN-COV could be used to prevent Covid-19 among persons with ongoing exposure to a SARS-CoV-2– infected individual. A household contact study design was utilized to assess whether REGEN-COV could prevent SARS-CoV-2 infection in a scenario with high risk of lateral transmission; this scenario was considered generalizable to other prevention settings. Here, we report the primary results of the phase 3 trial in adults and adolescents.

## METHODS

### Trial Design

This randomized, double-blind, placebo-controlled, two-part, phase 3 trial assessed the efficacy and safety of subcutaneous REGEN-COV in (Part A) preventing SARS-CoV-2 infection among uninfected household contacts of infected individuals and (Part B) also in treating recently infected asymptomatic patients (ClinicalTrials.gov number, NCT04452318). The trial was conducted at 112 sites in the United States (US), Romania, and Moldova. The trial is managed jointly by Regeneron, the Covid-19 Prevention Network (CoVPN), and the National Institute of Allergy and Infectious Diseases (NIAID).

Nasopharyngeal and serum samples were collected at the screening/baseline visit for central lab RT-qPCR testing and serum antibody testing. RT-qPCR was used to determine ongoing infection with SARS-CoV-2, while serology (anti-spike [S1] IgA, anti-spike [S1] IgG, and/or anti-nucleocapsid IgG) determined a prior or ongoing infection in which an innate antibody immune response had already occurred (i.e., seropositive; as opposed to seronegative). Part A included those who were RT-qPCR–negative, and Part B included those who were RT-qPCR–positive. The populations for Parts A and B were mutually exclusive and analyzed separately with different hierarchies and separate alpha allocation. Here we describe results for Part A.

Study participants were randomized (1:1) to receive REGEN-COV 1200 mg (600 mg each of casirivimab and imdevimab) or placebo via subcutaneous injection and were stratified by SARS-CoV-2 local diagnostic results and age. The trial consisted of a 1-day screening/baseline period, a 28-day efficacy assessment period (EAP), and a 7-month follow-up period (**Figure S1**). Enrollment was gated by sentinel safety review of the first 30 adults and 12 adolescents. The protocol is available upon request.

### Trial Oversight

Details are provided in the **Supplementary Appendix**.

### Study Participants

Asymptomatic, heathy adult (≥18 years of age) and adolescent (≥12 to <18 years) household contacts of the first known household member with SARS-CoV-2 infection (defined as the index case) were eligible if they anticipated living with the index case for at least 28 days. Participants were randomized within 96 hours of collection of the index case’s positive SARS-CoV-2 result, and individuals with prior SARS-CoV-2 infection were excluded. Full inclusion and exclusion criteria are provided in the **Supplementary Appendix**.

### Intervention and Assessments

At baseline (day 1), participants received 1200 mg REGEN-COV or placebo, administered as 4 subcutaneous injections in the abdomen (and thighs, as necessary), each containing 2.5 mL of active drug supplied as a 120 mg/mL solution or matching placebo. Signs and symptoms of Covid-19 were collected weekly by investigator interview about adverse events since the last visit/contact. If a participant became SARS-CoV-2 RT-qPCR–positive, signs and symptoms (type and severity) were collected weekly until resolution.

Nasopharyngeal swabs were collected for SARS-CoV-2 RT-qPCR at baseline and weekly during the EAP. For individuals testing positive, weekly swabs were collected until they tested negative twice. Analytical methods have been previously described.^7^

Some participants were referred to this study through the index case’s participation in a sister study of intravenous treatment with REGEN-COV or placebo in outpatients with Covid-19, COV-2067 (ClinicalTrials.gov number, NCT04425629). Participants were interviewed at baseline to collect information on household members, including the index case, and subject identification numbers between the two studies were linked so that it could be determined whether treatment of an index case impacted transmission among household contacts.

### Endpoints

The prespecified Part A primary efficacy analysis population consisted of participants without evidence of prior infection (RT-qPCR–negative/seronegative) randomized by January 28, 2021, excluding participants from the initial descriptive assessment (described in the **Supplementary Appendix**). The primary efficacy endpoint was the proportion of individuals with symptomatic, RT-qPCR–confirmed SARS-CoV-2 infection during the 28-day EAP. A broad-term definition of what constituted symptomatic Covid-19 was utilized for this analysis. Alternative definitions of symptomatic disease (strict-term and CDC) were also used for secondary analyses and are detailed in the **Supplementary Appendix**. Primary and key secondary endpoints were tested hierarchically (**Table S1**). Full lists of secondary efficacy and exploratory endpoints are provided in the statistical analysis plan.

Safety endpoints included treatment-emergent adverse events (TEAEs) and adverse events of special interest (AESIs): grade ≥3 hypersensitivity or injection site reactions. Safety endpoints are reported for participants randomized through January 28, 2021, until the data cut-off date of March 11, 2021, including those in the initial descriptive assessment.

### Statistical Analysis

The statistical analysis plan was finalized prior to database lock and treatment unblinding. The seronegative modified full analysis set included all randomized participants 12 years of age who were confirmed by central laboratory testing to be negative for SARS-CoV-2 by RT-qPCR (Part A) serology at baseline, excluding participants from the initial descriptive assessment. All efficacy endpoints are reported through the 28-day EAP. Safety data are reported for all participants who received study drug, including those in the initial descriptive assessment.

Simulations showed that approximately 1248 seronegative participants from 430 households (assuming an average household size of 2.9 participants) would provide >90% power to detect a relative risk of 0.5 (50% risk reduction of the assumed 10% attack rate in the placebo group), equivalent to an odds ratio of 0.47 at a two-sided alpha of 0.05.

Since the proportion of households with only a single study participant in the primary analysis population was more than 80%, the primary efficacy endpoint was analyzed using logistic regression. The model included fixed category effects of treatment group (placebo vs. REGEN-COV), region (US vs. ex-US), and age (≥12 to <50 years, ≥50 years of age). Analyses of key secondary efficacy endpoints are described in the **Supplementary Appendix**. A statistical hierarchy was employed to control for type 1 error based on a two-sided alpha of 0.5 to test primary and key secondary endpoints. Details regarding missing data imputation are provided in the **Supplementary Appendix** and the statistical analysis plan.

The population and methods for the pharmacokinetic analyses are described in the **Supplementary Appendix**.

## RESULTS

### Demographics and Baseline Characteristics

This study included 2475 randomized participants, not including those in the initial descriptive assessment, of whom 2067 (83.5%) were confirmed SARS-CoV-2 RT-qPCR–negative (and were analyzed as Part A), of which 1505 participants (72.8%) also showed no evidence of prior SARS-CoV-2 infection by serology testing (seronegative at baseline). These 1505 participants, without evidence of prior or ongoing infection (primary efficacy analysis population), were assigned to receive REGEN-COV (n=753) or placebo (n=752), respectively (**Figure S2**).

Mean age was 42.9 years, 45.9% were male, 9.3% identified as Black or African American, and 40.5% identified as Hispanic or Latino. Four-hundred and fifty-nine (30.5%) participants had a risk factor for Covid-19. The median (IQR) household size, including the index case and other household members who were not required to participate in the study, was 3 (2). The proportion of households consisting of only one RT-qPCR–negative (Part A), seronegative participant was 81.8% (**Table 1**).

**Table 1.** Demographics and Baseline Characteristics (Seronegative).*

|  | Placebo<br>(N=752) | REGEN-COV<br>1200 mg SC<br>(N=753) | Total<br>(N=1505) |
| --- | --- | --- | --- |
| Age — yr |  |  |  |
| Mean (range) | 42.7 (12–92) | 43.2 (12–87) | 42.9 (12–92) |
| ≥50 — no. (%) | 280 (37.2) | 294 (39.0) | 574 (38.1) |
| Sex — no. (%) | 358 (47.6) | 333 (44.2) | 691 (45.9) |
| Race — no. (%) |  |  |  |
| White | 635 (84.4) | 653 (86.7) | 1288 (85.6) |
| Black or African American | 78 (10.4) | 62 (8.2) | 140 (9.3) |
| Asian | 19 (2.5) | 23 (3.1) | 42 (2.8) |
| American Indian or Alaska Native | 4 (0.5) | 3 (0.4) | 7 (0.5) |
| Native Hawaiian or Pacific Islander | 2 (0.3) | 1 (0.1) | 3 (0.2) |
| Other | 14 (1.9) | 11 (1.5) | 25 (1.7) |
| Ethnicity — no. (%) |  |  |  |
| Hispanic or Latino | 319 (42.4) | 291 (38.6) | 610 (40.5) |
| Not Hispanic or Latino | 428 (56.9) | 459 (61.0) | 887 (58.9) |
| Other | 5 (0.7) | 3 (0.4) | 8 (0.5) |
| Mean weight — kg | 81.2±19.72 | 81.3±19.92 | 81.3±19.81 |
| Body-mass index† |  |  |  |
| Mean | 28.5±6.28 | 28.9±12.35 | 28.7±9.79 |
| >30 — no. (%) | 243 (32.3) | 260 (34.5) | 503 (33.4) |
| Participants with any high-risk factor for Covid-19 — no. (%) | 221 (29.4) | 238 (31.6) | 459 (30.5) |
| ≥65 years of age | 55 (7.3) | 76 (10.1) | 131 (8.7) |
| Body-mass index† ≥35 kg/m<sup>2</sup> | 104 (13.8) | 99 (13.1) | 203 (13.5) |
| Chronic kidney disease | 11 (1.5) | 17 (2.3) | 28 (1.9) |
| Diabetes | 45 (6.0) | 58 (7.7) | 103 (6.8) |
| Immunosuppressive disease | 2 (0.3) | 5 (0.7) | 7 (0.5) |
| Receiving immunosuppressive treatment | 11 (1.5) | 4 (0.5) | 15 (1.0) |
| ≥55 years of age with CVD, hypertension, or COPD | 90 (12.0) | 99 (13.1) | 189 (12.6) |
| Total no. of households | 686 | 679 | 1209 |
| Number of households by size — no. (%)‡ |  |  |  |
| 1 | 503 (73.3) | 486 (71.6) | 989 (81.8) |
| 2 | 136 (19.8) | 146 (21.5) | 172 (14.2) |
| 3 | 30 (4.4) | 30 (4.4) | 31 (2.6) |
| 4 | 13 (1.9) | 13 (1.9) | 13 (1.1) |
| >4 | 4 (0.6) | 4 (0.6) | 4 (0.3) |
| Participants with an index case participating in study COV-2067<br>— no. (%) | 186 (24.7) | 187 (24.8) | 373 (24.8) |

\*Plus-minus values are means ± SD. COPD denotes chronic obstructive pulmonary disease, CVD cardiovascular disease, and SC subcutaneous.
†The body-mass index is the weight in kilograms divided by the square of the height in meters.
‡Household size is calculated by counting the seronegative study participants in Part A. Percentages are based on the number of households as the denominator, instead of the number of participants.

Approximately 25% of participants lived with an index case that was receiving REGEN-COV or placebo in a sister study involving outpatients with symptomatic Covid-19, COV-2067 (**Table 1**). Treating index cases in COV-2067 had no impact on household contacts in this study; these results are described in the **Supplementary Appendix**.

Demographics and baseline characteristics for seropositive participants are presented in **Table S2**.

### Efficacy

#### Prevention of SARS-CoV-2 Infection

Subcutaneous REGEN-COV significantly prevented symptomatic SARS-CoV-2 infection compared with placebo (81.4% risk reduction; 11/753 [1.5%] vs. 59/752 [7.8%], respectively; odds ratio [OR] 0.17; P<0.0001; **Table 2**); efficacy was apparent within days of treatment (**Figure 1A**). REGEN-COV prevented 71.9% of infections within the first week (9/753 [1.2%] vs. 32/752 [4.3%] for REGEN-COV vs. placebo, respectively), and 92.6% of infections in weeks 2–4 (2/753 [0.3%] vs. 27/752 [3.6%] for REGEN-COV vs. placebo, respectively; post-hoc analysis; **Table S3**). Findings were similar when utilizing broad-term, strict-term, and CDC definitions of symptomatic SARS-CoV-2 infection (**Table S4**) and regardless of baseline serology status (**Table S5**).

**Table 2.** Primary and Key Secondary Efficacy Endpoints.*

|  | **Placebo**<br>**(N=752)** | **REGEN-COV**<br>**1200 mg SC**<br>**(N=753)** |
| --- | --- | --- |
| Proportion of participants who have a symptomatic RT-qPCR–confirmed SARS-CoV-2 infection (broad-term)† |  |  |
| no./total no. (%) | 59/752 (7.8) | 11/753 (1.5) |
| Relative risk reduction | - | 81.4% |
| Odds ratio (95% CI) | - | 0.17 (0.09–0.33) |
| P value¶ | - | <0.0001 |
| Proportion of all participants with viral load >10<sup>4</sup> copies/ml□ |  |  |
| no./total no. (%) | 85/749 (11.3) | 12/745 (1.6) |
| Relative risk reduction | - | 85.8% |
| Odds ratio (95% CI) | - | 0.13 (0.07–0.24) |
| P value¶ | - | <0.0001 |
| No. of weeks of symptomatic RT-qPCR–confirmed SARS-CoV-2 infection (broad-term) |  |  |
| Total no. of weeks | 187.7 | 12.9 |
| Total duration (weeks) per 1000 participants | 249.6 | 17.1 |
| Reduction□ | - | 93.1% |
| P value§ | - | <0.0001 |
| Mean per-symptomatic infected participant duration of symptoms — weeks | 3.2±2.68 | 1.2±0.99 |
| No. of weeks of high viral load (>10<sup>4</sup> copies/ml) among all participants□ |  |  |
| Total no. of weeks | 136.0 | 14.0 |
| Total duration (weeks) of per 1000 participants | 181.6 | 18.8 |
| Reduction□ | - | 89.6% |
| P value§ | - | <0.0001 |
| Mean per-infected participant duration of high viral load — weeks | 1.3±0.87 | 0.4±0.60 |
| No. of weeks of any RT-qPCR–confirmed SARS-CoV-2 infection (symptomatic or asymptomatic) |  |  |
| Total no. of weeks | 231.0 | 41.0 |
| Total duration (weeks) per 1000 participants | 307.2 | 54.4 |
| Reduction□ | - | 82.3% |
| P value§ | - | <0.0001 |
| Mean per-infected participant duration of overall infection — weeks | 2.2±1.07 | 1.1±0.42 |
| Proportion of participants who have any RT-qPCR–confirmed SARS-CoV-2 infection (symptomatic or asymptomatic) |  |  |
| no./total no. (%) | 107/752 (14.2) | 36/753 (4.8) |
| Relative risk reduction | - | 66.4% |
| Odds ratio (95% CI) | - | 0.31 (0.21–0.46) |
| P value# | - | <0.0001 |

\*Plus–minus values are means ± SD. Key secondary endpoints are presented in order of the hierarchy testing sequence. RT-qPCR denotes quantitative reverse transcription polymerase chain reaction, SC subcutaneous.
†Primary endpoint
□ For viral load endpoints, only participants with at least one viral load assessment during the 28-day efficacy assessment period are included for analysis.
§Based on a stratified Wilcoxon rank sum test (van Elteren test) with region (US vs. ex-US) and age group (12 to <50 years of age vs. ≥ 50 years of age) as strata.
□ Based on the normalized weeks per 1000 participants.
¶Based on logistic regression model adjusted by region (US vs. ex-US) and age group (12 to <50 years of age vs. ≥ 50 years of age).

**Figure 1.**
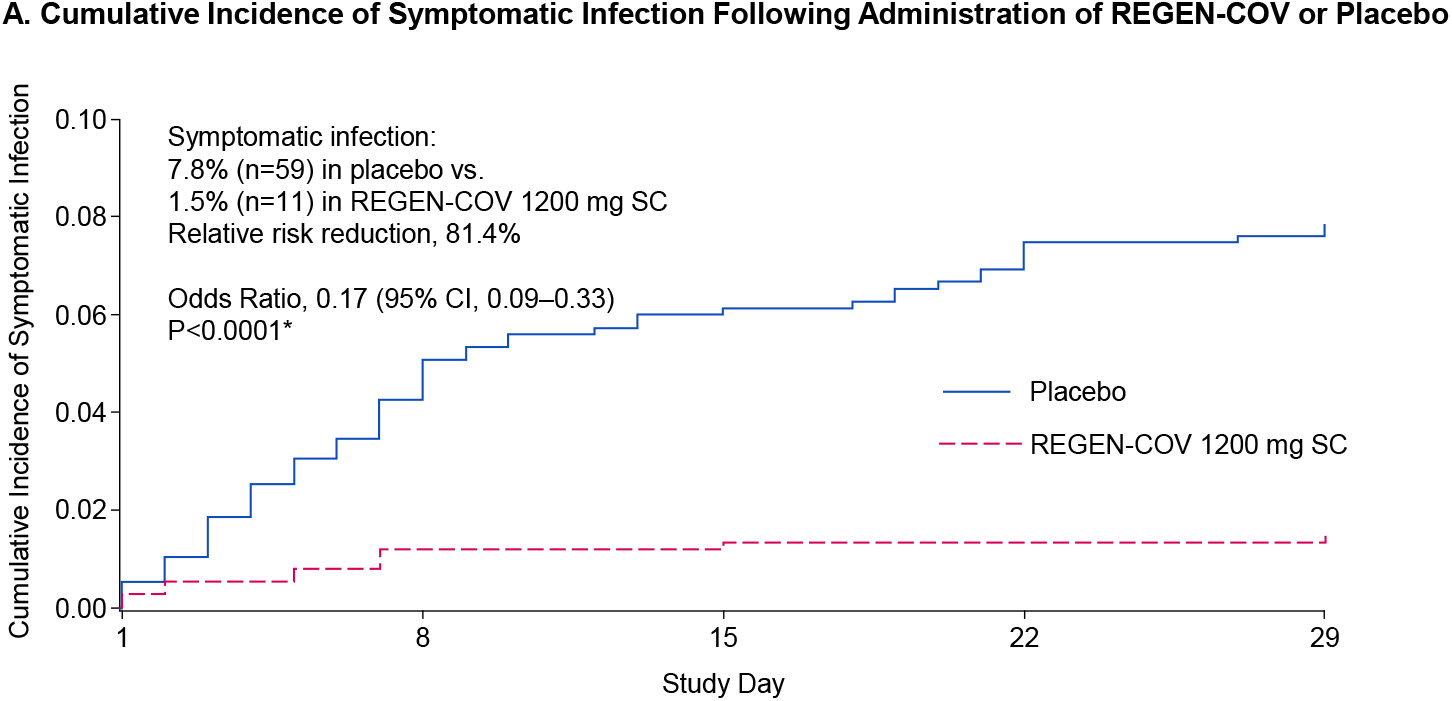
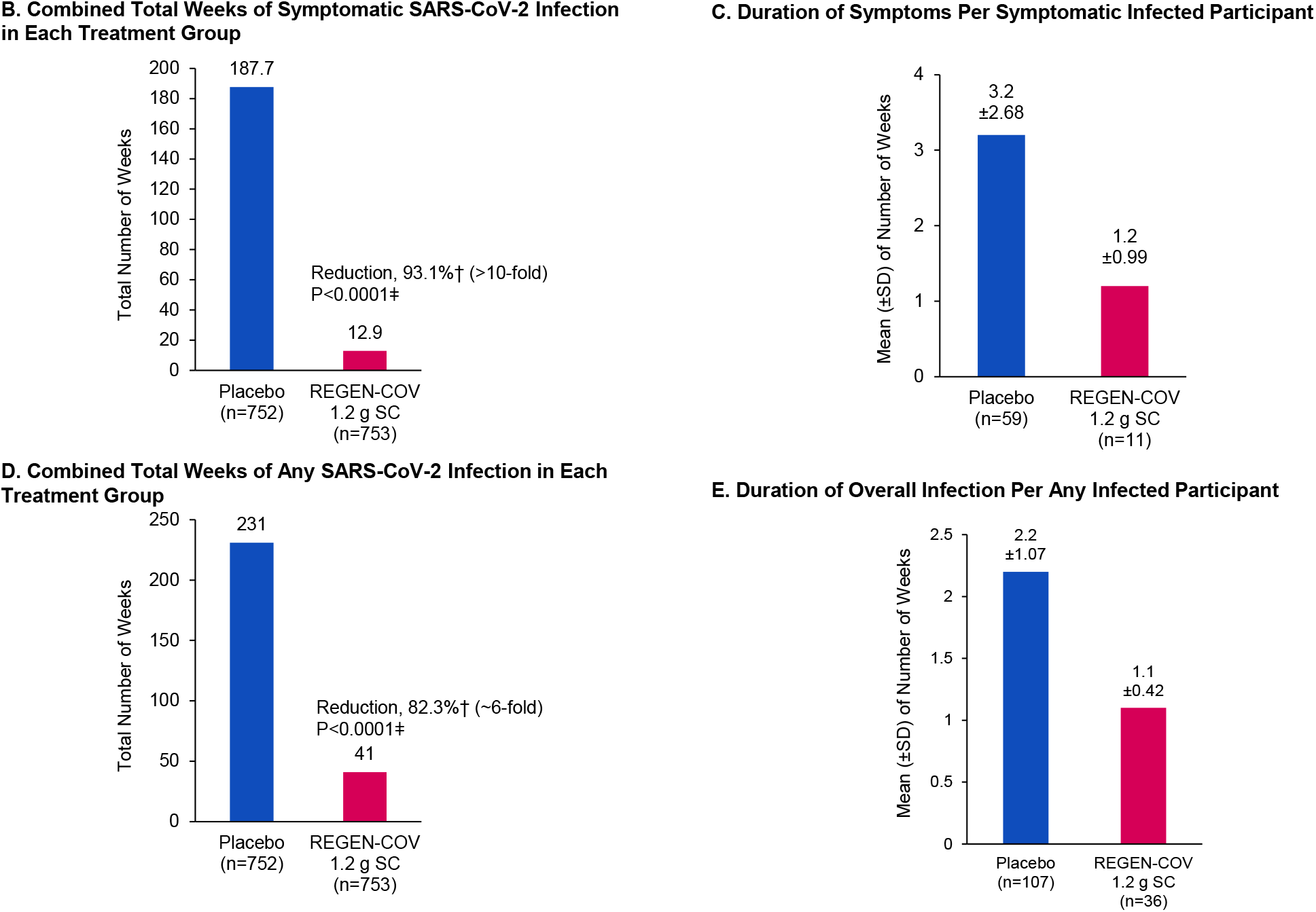
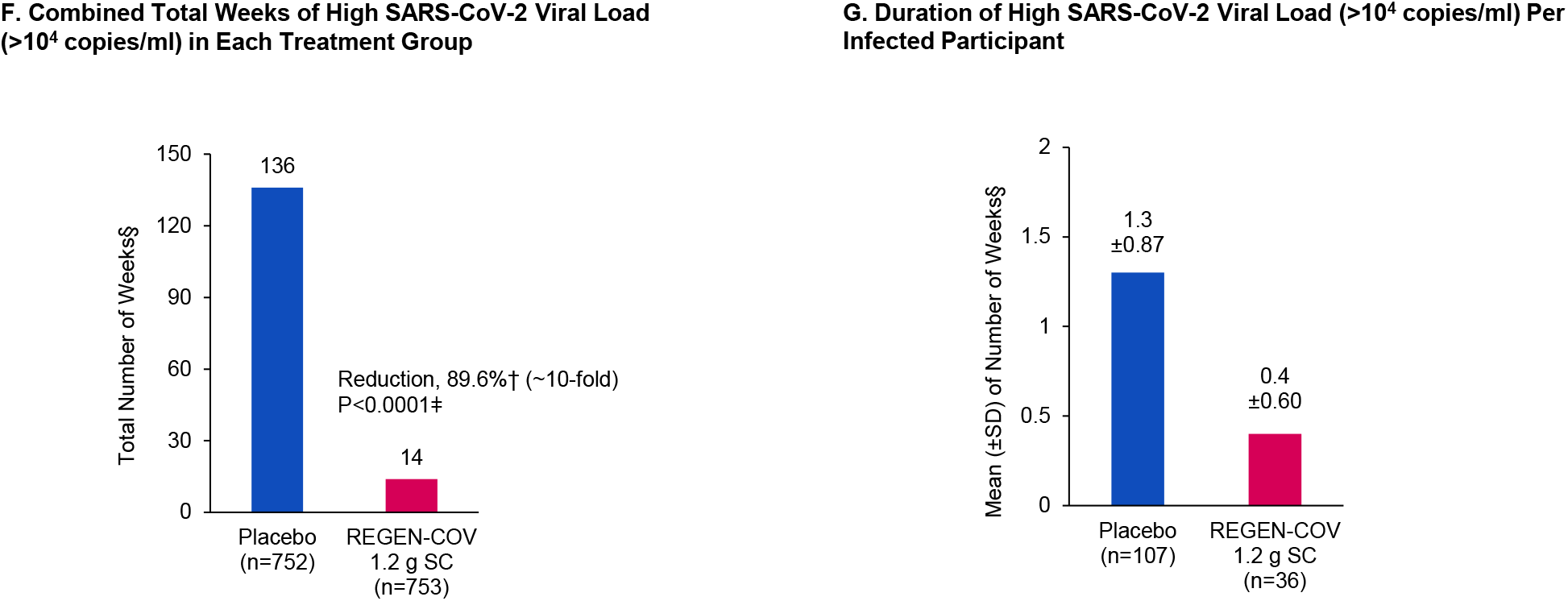
REGEN-COV Reduces Symptomatic Infection in Those Who Are Uninfected at Baseline. Panel A shows cumulative incidence of symptomatic infection following administration of REGEN-COV or placebo. Panel B shows combined total weeks of symptomatic SARS-CoV-2 infection in each treatment group. Panel C shows duration of symptoms per symptomatic infected participant. Panel D shows combined total weeks of any SARS-CoV-2 infection in each treatment group. Panel E shows duration of overall infection per any infected participant. Panel F shows combined total weeks of high SARS-CoV-2 viral load (>10^4^ copies/ml) in each treatment group. Panel G shows duration of high SARS-CoV-2 viral load (>10^4^ copies/ml) per infected participant. *Based on a logistic regression model including fixed category effects of treatment group (placebo vs. REGEN-COV), region (US vs. ex-US), and age (≥12 to <50 years of age, ≥50 years of age). †Based on the normalized weeks per 1000 participants. ǂBased on a stratified Wilcoxon rank sum test (van Elteren test) with region (US vs. ex-US) and age group (12 to <50 years of age vs. ≥50 years of age) as strata. §If a visit had missing viral load data, that visit was not included in the analysis. Only participants with at least one post-baseline viral load nasopharyngeal swab samples were included in this analysis. CI denotes confidence interval, SARS-CoV-2 severe acute respiratory syndrome coronavirus 2, SC subcutaneous, and SD standard deviation.

There was a 93.1% reduction in the aggregated total number of weeks with symptoms with REGEN-COV vs. placebo: 12.9 weeks vs. 187.7 weeks, respectively; P<0.0001 (**Figure 1B**; **Table 2**). This corresponded to a 2-week reduction in the mean duration of infection per symptomatic participant from 3.2 weeks in the placebo group to 1.2 weeks in the REGEN-COV group (**Figure 1C**; **Table 2**).

Subcutaneous REGEN-COV significantly prevented overall SARS-CoV-2 infections (asymptomatic and symptomatic) compared with placebo (66.4% risk reduction; 36/753 [4.8%] vs. 107/752 [14.2%], respectively; OR 0.31; P<0.0001; **Table 2**). Consistent with this finding, there was an 82.3% reduction in the aggregated total number of weeks of any RT-qPCR–detectable infection with REGEN-COV vs. placebo: 41.0 weeks vs.

231.0 weeks, respectively; P<0.0001 (**Figure 1D**; **Table 2**). This corresponded to an approximate 1-week reduction per any infected participant in the mean duration of overall infection from 2.2 weeks in the placebo group to 1.1 weeks in the REGEN-COV group (**Figure 1E**; **Table 2**).

Additionally, there was an 85.8% risk reduction in the proportion of all participants with high SARS-CoV-2 viral load, defined as >10^4^ copies/mL via nasopharyngeal RT-qPCR, with REGEN-COV vs. placebo: 12/745 (1.6%) vs. 85/749 (11.3%), respectively; OR 0.13; P<0.0001 (**Table 2**). Of participants who became infected after REGEN-COV treatment, the majority had low viral load (**Table S6**). Consistent with this finding, there was an 89.6% reduction in the total number of weeks of high SARS-CoV-2 viral load with REGEN-COV treatment vs. placebo: 14.0 weeks vs. 136.0 weeks, respectively; P<0.0001 (**Figure 1F**; **Table 2**). This corresponded to an approximate 6-day reduction per infected participant in the mean duration of high viral load infection from 1.3 weeks in the placebo group to 0.4 weeks in the REGEN-COV group (**Figure 1G**; **Table 2**).

Participants who became infected despite REGEN-COV treatment also exhibited decreased peak viral load compared to infected individuals in the placebo group (**Figure 2A**) and had a shorter duration of viral RNA detection (viral load infections >10^4^ copies/mL; **Figure S3**). REGEN-COV treatment prevented high viral load levels in both symptomatic and asymptomatic participants (**Figure 2B and 2C**). Additional viral load data is shown in **Table S7**.

**Figure 2.**
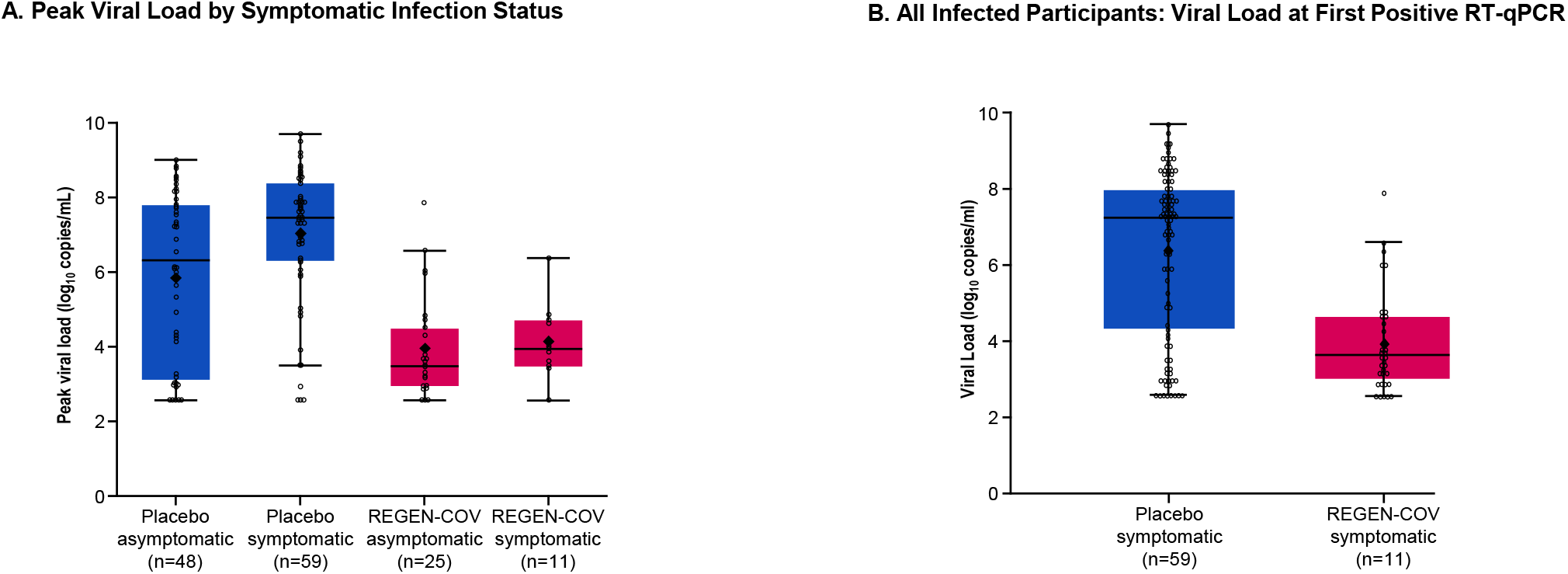
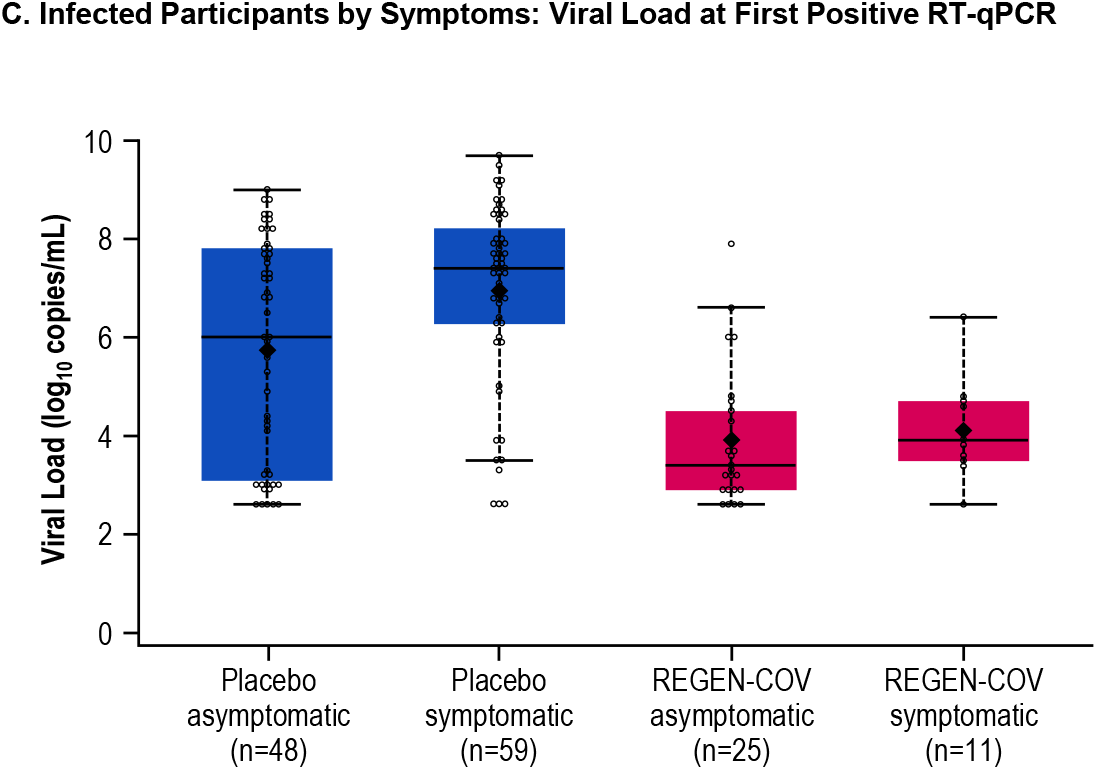
Individuals Who Become Infected Despite REGEN-COV Treatment Demonstrate Lower Viral Burden Compared With Placebo-Treated Individuals. Panel A shows peak viral load by symptomatic infection status. Panel B shows all infected participants: viral load at first positive RT-qPCR. Panel C shows infected participants by symptoms: viral load at first positive RT-qPCR. Lines in the boxes represent the median. Large, bolded dots in the boxes represent the mean. Bottom and top of boxes represent quartiles 1 (25th percentile) and 3 (75th percentile), respectively. Whiskers represent the 1.5 times interquartile range. RT-qPCR denotes quantitative real-time polymerase chain reaction, and SC subcutaneous.

#### Sub-analyses in Adolescents Ages 12-17

In adolescent participants, the rate of symptomatic SARS-CoV-2 infection was 0% (0/34) in the REGEN-COV group compared with 11.8% (4/34) in the placebo group, corresponding to a risk reduction of 100%. REGEN-COV was associated with a 100% reduction in the risk of symptomatic infection regardless of serology status: 0% (0/46) in the REGEN-COV group compared with 9.3% (4/43) in the placebo group.

### Safety

REGEN-COV was generally well tolerated. The proportion of participants in the REGEN-COV and placebo groups who experienced at least one TEAE was 20.2% vs. 29.0% overall, respectively, and 16.0% vs. 16.5% for non–Covid-19 TEAEs, respectively (**Table S8**). TEAEs occurring in ≥2% of participants included Covid-19, asymptomatic Covid-19, headache, and injection site reaction (**Table 3**). No AESIs were reported during the study, and no participants withdrew from the study due to an adverse event.

**Table 3.** Treatment-Emergent Adverse Events Occurring in ≥2% of Participants.*

| **Preferred Term — no. of participants (%)** | **Placebo**<br>**(N=1306)** | **REGEN-COV**<br>**1200 mg SC**<br>**(N=1311)** |
| --- | --- | --- |
| Covid-19 | 112 (8.6) | 15 (1.1) |
| Asymptomatic Covid-19 | 108 (8.3) | 54 (4.1) |
| Headache | 46 (3.5) | 24 (1.8) |
| Injection site reaction | 19 (1.5) | 55 (4.2) |

\*Regardless of the SARS-CoV-2 serology status at baseline. SC denotes subcutaneous.

Serious adverse events (SAEs) were experienced by 0.8% and 1.1% of participants in the REGEN-COV and placebo groups, respectively (**Table S9**). None of the SAEs in the REGEN-COV group were considered to be Covid-19– or drug-related. There were no participants with emergency room (ER) visits or hospitalizations due to Covid-19 in the REGEN-COV group, whereas there were 4 participants in the placebo group. Two deaths (0.2%) occurred in each treatment group during the study outside of the EAP, none of which were attributed to Covid-19 (**Table S10**). In the REGEN-COV group, one participant died of congestive cardiac failure, and one participant who had multiple comorbidities experienced sudden death not related to Covid-19. In the placebo group, one participant died of a gunshot wound, and one participant died of cardiac arrest not related to Covid-19.

### Pharmacokinetics

Casirivimab and imdevimab were rapidly absorbed (**Figure S4**), with mean concentrations in serum one day after dosing of 22.1 mg/L and 25.8 mg/L, respectively. The antibodies reached maximal concentrations in serum at a median time of 7 to 8 days. Casirivimab and imdevimab exhibited linear elimination and had mean half-lives of 32.4 days and 27.0 days, respectively. Mean concentrations in serum 28 days after dosing were 30.4 mg/L for casirivimab and 24.6 mg/L for imdevimab, levels above the estimated target dose for neutralization of SARS-CoV-2 (20 mg/L). A summary of pharmacokinetic parameters is shown in **Table S11**.

## DISCUSSION

More than 173 million individuals have been infected with SARS-CoV-2 worldwide, and more than 3.7 million died from complications from Covid-19.^9^ At the time we initiated the REGEN-COV program, we hypothesized that a combination of neutralizing, non-competing, monoclonal antibodies could have activity in both treatment and prevention of SARS-CoV-2 infection while retaining activity against the inevitable emergence of viral variants. In the outpatient setting, treatment with intravenous REGN-COV reduced Covid-19–related hospitalization or all-cause death, rapidly resolved symptoms, and reduced viral load.^7,8^

In designing this phase 3 clinical trial, we anticipated a high background rate of lateral transmission in a household with a documented SARS-CoV-2–infected individual quarantining alongside other household members, as has subsequently been confirmed.^10^ This scenario serves as an exemplar for many other scenarios where uninfected individuals are in close proximity and would be at high risk for infection, including hospitals, nursing homes, dense housing complexes, and schools. Prevention in high-risk settings should also correlate with prevention in lower risk settings.

Among household members who were confirmed to be both uninfected and without recent history infection, subcutaneous REGEN-COV prevented both symptomatic and overall SARS-CoV-2 infections and was well tolerated. Subcutaneous REGEN-COV substantially and significantly prevented symptomatic SARS-CoV-2 infection by ∼81%: REGEN-COV significantly prevented infections within one week of dosing (∼72% reduction), which increased (∼93% reduction) after the first week. REGEN-COV also reduced high viral load infections (>10^4^ copies/mL) by ∼86% and all infections (symptomatic and asymptomatic) by ∼66%. Individuals who still developed SARS-CoV-2 infection despite receiving REGEN-COV had a lower likelihood of developing symptoms. In the minority of participants who became infected after REGEN-COV treatment and developed laboratory-confirmed symptomatic infection (1.5%), the duration of infection was reduced by 2 weeks. Moreover, in participants who developed either symptomatic or asymptomatic infection despite REGEN-COV treatment, the magnitude and duration of detectable RNA (weeks of RT-qPCR positivity and peak viral load) were markedly decreased. A heat map showing presence of symptoms and viral load over time in infected individuals shows the prolonged time course for individuals receiving placebo compared with REGEN-COV–treated individuals (**Figure S3**).

The incidence of TEAEs was higher in the placebo group than the group receiving REGEN-COV, with the difference attributed to the higher number of Covid-19 infections observed in the placebo group. No grade 3 or higher injection site or hypersensitivity reactions occurred. No individuals receiving REGEN-COV ended up with an ER visit or hospitalization during the EAP, compared with 4 individuals receiving placebo. Following subcutaneous dosing, concentrations of each antibody in serum were well above the predicted neutralization target concentration (based preclinical data) as early as the first day following dosing and throughout the 28-day EAP.

While the primary efficacy analysis was in seronegative individuals, when analyses were conducted irrespective of baseline serological status, REGEN-COV also substantially prevented symptomatic infection (**Table S5**). Combined with the observed tolerability of REGEN-COV, this suggests that point of care serology data will not be needed for treatment decisions in the clinic.

These data support the potential use of REGEN-COV to prevent SARS-CoV-2 infection and symptomatic disease in individuals who require immediate protection; REGEN-COV treatment of such individuals could decrease further spread and transmissibility. Importantly, REGEN-COV maintains its activity against emerging variants of concern.^4-6^ This trial also demonstrated that subcutaneous administration of REGEN-COV is efficacious with an acceptable safety profile, thus potentially providing substantial benefits by avoiding the healthcare resources necessary for an intravenous infusion.

Despite increasing utilization of highly effective vaccines, we are a long way from eradicating SARS-CoV-2. Moreover, we do not yet understand how many people will ultimately choose to become vaccinated, how vaccine efficacy will wane over time, nor the impact of emerging variants of concern. For these reasons, there will long remain a need for a complementary approach to prevent spread of SARS-CoV-2 in populations 1) who are still not vaccinated, 2) who have waning vaccine mediated protection due to time or the emergence of variants, or 3) who are immunocompromised and cannot mount an antibody-mediated immune response, who remain at risk due to exposure to ongoing viral reservoirs. Millions in the US alone suffer from either a primary or secondary immune deficiency; recent studies show that the majority of such patients mount insufficient response to vaccines.^11-13^ Additionally, immunosenescence may compromise immune response following vaccination.^14,15^ Such immunocompromised individuals will require an alternative chronic prophylaxis approach until SARS-CoV-2 has been substantially eradicated. This study showed that throughout the 28-day observation period, the achieved concentrations of a single subcutaneous dose of REGEN-COV prevented symptomatic infection, suggesting a potential for REGEN-COV to be used for chronic prophylaxis in individuals at risk for SARS-CoV-2 infection.

In conclusion, a single subcutaneous administration of 1200 mg of REGEN-COV dramatically reduced symptomatic and asymptomatic infection over the 28-day assessment period in uninfected individuals at high risk for SARS-CoV-2 infection, with an acceptable safety profile. REGEN-COV should also be considered to rapidly protect at-risk, unvaccinated individuals or those who remain susceptible to SARS-CoV-2 infection despite vaccination.

## Supporting information

DB NIH Publishing Agreement

MM NIH Publishing Agreement

AB ICMJE

AM ICMJE

ATD ICMJE

ATH ICMJE

BJ ICMJE

BK ICMJE

CAK ICMJE

CBH ICMJE

DHB ICMJE

DMW ICMJE

DR ICMJE

DRB ICMJE

EF-N ICMJE

FI ICMJE

GDY ICMJE

GH ICMJE

IH ICMJE

JDD ICMJE

JDH ICMJE

JK ICMJE

K-CC ICMJE

KCT ICMJE

KJB ICMJE

LAP ICMJE

LL ICMJE

MAM ICMJE

MPO ICMJE

MSC ICMJE

NB ICMJE

NSarkar ICMJE

NStahl ICMJE

PH ICMJE

RM ICMJE

RP ICMJE

RV ICMJE

YK ICMJE

## Data Availability

Qualified researchers may request access to study documents (including the clinical study report, study protocol with any amendments, blank case report form, statistical analysis plan) that support the methods and findings reported in this manuscript. Individual anonymized participant data will be considered for sharing once the indication has been approved by a regulatory body, if there is legal authority to share the data and there is not a reasonable likelihood of participant re-identification. Submit requests to https://vivli.org/.

## DATA SHARING

A data sharing statement provided by the authors is available with the full text of this article.

## SUPPORTED BY

Supported by Regeneron Pharmaceuticals, Inc. and F. Hoffmann-La Roche Ltd. This trial was conducted jointly with the National Institute of Allergy and Infectious Diseases (NIAID), National Institutes of Health (NIH). The CoVPN is supported by cooperative agreement awards from NIAID, NIH. The content of this manuscript is solely the responsibility of the authors and does not necessarily represent the official views of the NIH. The work on this study was supported by CoVPN award number UM1AI068619.

## ACKNOWLEDGEMENT

We thank the study participants; their families; the investigational site members involved in this trial (listed in the **Supplementary Appendix**); the Covid-19 Phase 3 Prevention Trial Team (listed in the **Supplementary Appendix**); the members of the Data and Safety Monitoring Board; Caryn Trbovic, Ph.D., Brian Head, Ph.D., and S. Balachandra Dass, Ph.D., from Regeneron Pharmaceuticals for assistance with development of the manuscript; and Prime, Knutsford, UK, for formatting and copy editing suggestions.

