## Supplementary Appendix for "Subcutaneous REGEN-COV Antibody Combination for Covid-19 Prevention"

#### Covid-19 Phase 3 Prevention Trial Team (REGN 2069/CoVPN 3502)

##### Study Sites and Investigators

**Advanced Pulmonary Research Institute, Loxahatchee, FL:** Neal Warshoff, Liudmila Moreiras

**AGA Clinical Trials, Miami, FL:** Dario Altamirano, Dickson Ellington, Faisal Faikih

**AMR-Knoxville (formerly NOCCR), Knoxville, TN:** William Smith, Richard Gibson, Katie Buckner

**Ardmore Medical Research, Winston Salem, NC:** Robert Rosen, Amy Sapp

**Arizona Liver Health, Tucson, AZ:** Anita Kohli, Vicki McIntyre, Yessica Sachdeva

**Arizona Liver Health, Chandler, AZ:** Yessica Sachdeva, Anita Kohli, Amanda McFarland, Dina Gibson

**Ark Clinical Research, Long Beach, CA:** Kenneth Kim, Jason Ahn, Lisa Neinchel, Nayna Paryani, Amber Mottola, Eva Day, Martha Navarro

**Atella Clinical Research La Palma, CA:** Rafaelito Victoria, Xanthe Victoria, Rene Uong

**Atrium Health, Charlotte, NC:** Mindy Sampson, Christopher Polk, Michael Leonard, Lewis McCurdy, Leigh A. Medaris, Zainab Shahid, Lisa Davidson

**Avera McKennan Hospital and University Health Center, Sioux Falls, SD:** Jawad Nazir, John Lee, Amy Elliott, Swami Sathyanaryan, Mansi Oberoi, Muhammad (Danial) Siddiqui, Muhammad Arsad, Kara Bruning

**Adolescent & Young Adult Research at Core – NIAID CoVPN, Chicago, IL:** Sybil Hosek, Temitope Oyedele, Vanessa Sarda, Monica Mercon

**Beth Israel Deaconess Medical Center, Boston, MA:** Kathryn Stephenson, Dan Barouch, Boris Juelg, Chen Sabrina Tan, Rebecca Zash, Ai-ris Collier, Jessica Ansel, Kate Jaegle

**Bio-Medical Research, LLC, Miami, FL:** Lilia Roque-Guerrero, Ana Gomez Ramirez, Javier Capote, Gisel Paz

**Boston Medical Center Ped. HIV Program NICHD – NIAID CoVPN, Boston, MA:** Michael Paasche-Orlow, Julien Dedier

**California Medical Research Associates, Northridge, CA:** Sanjay Vadgama, Ramachandra Patak

**Cardiology Care Clinics, Eatonton, GA:** Nicolas Chronos, Cary Hefty

**Carolina Institute for Clinical Research, Fayetteville, NC:** Judith Borger, Ifeanyi Momodu, Lindsey Carswell, Benjamin King, Ryan Starr, Scott Syndergaard

**Carolina Medical Research, Clinton, SC:** Nancy Patel, Ravikumar Patel, Ryan Sattar

**Catalina Research Institute, Montclair, CA:** Rizwana Mohseni, Jeffrey Unger, Sheila De Jesus-Maranan, Cecilia Casaclang

**Centex Studies, Lake Charles, LA:** Michael Seep, Celeste Brown, Joshua Whatley

**Chicago Clinical Research Institute, Chicago, IL:** Dennis Levinson, Saad Alvi, Norman James, Azazuddin Ahmed,

**Clinical Research of Central Florida, Winter Haven, FL:** Robinson Koilpillai, Stephanie Cassady, Jennifer Cox, Eduardo Torres

**Clinical Trial of Florida LLC, Miami, FL:** James Krainson, Mark J. Rosenthal

**Crossroads Clinical Research, Corpus Christi, TX:** Michael Winnie, Jerry Plemons, Omesh Verma, Richard Leggett

**DM Clinical Research/BFHC Research, San Antonio, TX:** Ramon Reyes, Keith Beck, Brian Poliquin

**DM Clinical Research/LinQ Research, LLC, Pearland, TX:** Murtaza Mussaji, Jignesh Shah

**<u>East Coast Institute for Research, Jacksonville, FL</u>:** David Sutton, Edward Pereira, Rodel Gloria, Stacey Kelly, Amy Dennis-Saltz, Mae Sheikh-Ali, Elias Saikali, James Magee, Rebecca Goldfaden

**<u>Epic Medical Research, Red Oak, TX</u>:** Haresh Boghara, Sunny Patel, Bari Eichelbaum

**<u>Excel Clinical Research, Las Vegas, NV</u>:** Duane Anderson, Sean Su, Alexander Akhavan, Diana Kirby, Joy Venglik

**<u>Fenway Health – NIAID CoVPN, Boston, MA</u>:** Kenneth Mayer, Taimur Khan, Marcy Gelman

**<u>Florida Pulmonary Research Institute, LLC, Winter Park, FL</u>:** Faisal A. Fakih, Faisal M. Fakih, Daniel Layish, Fernando Alvarado, Jose Diaz

**<u>Fomat Medical Research, Oxnard, CA</u>:** Augusto Focil, Griselda Rosas, Stevan Correa, Michael Bogseth

**<u>Future Innovative Treatments, LLC, Colorado Springs, CO</u>:** Bhaktasharan Patel, Gary Tarshis, Katrina Grablin

**<u>Geisinger Medical Center, Danville, PA</u>:** Paul Simonelli, Stanley Martin, Alvin Sharma, Anna Chen, Pragya Dhaubhadel, Shaeesta Khan, Sreelatha Naik, Sudheer Penupolu, Thulashie Sivarajah, Tae-Sung Kwon, Lakshmi Saladi

**<u>Geisinger Wyoming Valley, Wilkes-Barre, PA</u>:** Paul Simonelli, Stanley Martin, Alvin Sharma, Anna Chen, Pragya Dhaubhadel, Shaeesta Khan, Sreelatha Naik, Sudheer Penupolu, Thulashie Sivarajah, Tae-Sung Kwon, Lakshmi Saladi

**<u>Harlem Hospital Center – New York City Health and Hospitals Corporation, Harlem, NY</u>:** Farbod Raiszadeh, Sharon Mannheimer, Khaing Myint, Hussein Assallum, Lovelyamma Varghese, Akari Kyawa

**<u>Harlem Prevention Center – NIAID CoVPN, Harlem, NY</u>:** Ellen Morrison, Sharon Mannheimer, Julie Franks, Jun Avelino Loquere, Orlando Rosario, Andrea Low, Joan Villacrucis

**<u>HD Research Group, Houston, TX</u>:** Alan Skolnick, Harold Minkowitz, David Leiman, Todd Price, Anatoli Krasko, Idisoro Wiener

**Healthcare Research Network, Hazelwood, MO:** Larry Reed, Oscar Lin

**Henry Ford Health System, Detroit, MI:** Mayur Ramesh, George Alangaden

**Holy Name Medical Center, Teaneck, NJ:** Suraj Saggar, Thomas Birch, Benjamin De La Rosa, Karyna Neyra, Erina Kunwar

**IACT Health, Columbus, GA:** Jeffrey Kingsley, April Pixler, Veronica McBride

**Icahn School of Medicine at Mount Sinai, New York, NY:** Judith Aberg, Michelle Cespedes, Alexandra Abrams-Downey, Erna Kojic, Luz Lugo, Sean Liu, Nadim Salomon, David Perlman, Deena Altman, Farah Rahman, Georgina Osorio, Joseph Mathew, Sanjana Koshy, Dana Mazo, Francesca Cossarini, Sondra Middleton, Alina Jen, Erika Maria Reategui Schwarz

**Innovative Research of West Florida, Clearwater, FL:** Miguel Trevino, Benjamin DeVries

**Lincoln Medical Center – New York City Health and Hospitals Corporation, Bronx, NY:** Vidya Menon, Moiz Kasubhai, Usha Venugopal, Anjana Pillai, Franscene Oulds

**M3 Wake Research, Raleigh, NC:** Matthew Hong, Wayne Harper, Lynn Eckert, Douglas Wadeson, Lisa Cohen

**Maryland School of Medicine, Baltimore, MD:** Joel Chua, Shyam Kottilil, Jennifer Husson, John Baddley, R. Gentry Wilkerson, Shivakumar Narayanan, Uzoamaka Eke, Myint Noe, Melanie Malave Sanchez

**Massachusetts General Hospital – ID Clinical Research Unit – NIAID CoVPN, Boston, MA:** Arthur Kim, Greg Robbins, Mark Siedner, Rajesh Gandhi, Kristen Hysell, Jacob Lazarus, Lael Yonker

**McGovern Medical School at The University of Texas Health Science Center, Houston, TX:** Roberto Arduino, Karen J. Vigil

**Medical Research of Westchester, Miami, FL:** Richard Perez-Perez, Carlos J. Bello, Esperanza Arce-Nunez, Jorge Acosta, Julio L. Arronte

**Medical University of South Carolina, Charleston, SC:** Eric Meissner, Patrick Flume, Andrew Goodwin, Deeksha Jandhyala, Nandita Nadig

**MedPharmics, Metairie, LA:** Robert Jeanfreau, Susan Jeanfreau, Susan Tortorich, Shiva Akula

**MedPharmics, Gulfport, MS:** Paul Matherne, Donald Gaddy, Magdy Mikhail

**Mercury Clinical Research, Houston, TX:** Rajasekaran Annamalai, Huy Nguyen, Nizar Nayani, Mahalakshmi Ramchandra

**META Medical Research Institute, Dayton, OH:** Priyesh Mehta, Jacqueline Horne, Grace Hassan

**Midland Florida Clinical Research Center, Deland, FL:** Godson Oguchi, Judepatricks Onyema

**Midway Immunology and Research Center, Fort Pierce, FL:** Moti Ramgopal, Brenda Jacobs, Lisa Cason, Angela Trodglen

**National Institute of Infectious Diseases, Bucharest, Romania:** Adrian Streinu, Daniela Manolache, Anca Streinu-Cercel, Oana Sandulescu, Ana Blanaru, Monica Stoica, Ana Maria Andone, Daniela Dospinoiu, Silviu Serban, Loredana Patru, Christina Buhuara, Ramona Dorobantu, Magdalena Motoi, Ioana Daramus, George Bihoi, Alexandra Ghita, Victor Miron, Gylda Spataru

**New Jersey Medical School Clinical Research Center – NIAID CoVPN, Newark, NJ:** Amesika Nyaku, Shobha Swaminathan

**Next Level Urgent Care, Houston, TX:** Terence Chang, Robbyn Traylor, Lenee Gordon, John McDivitt, Lizette Castro

**Northern California Research, Sacramento, CA:** Douglas Young, Gary Carson

**New York University Langone Vaccine Center – NIAID CoVPN, Manhattan, NY:** Angelica Kottkamp, Mark J. Mulligan, Anna Bershteyn, Vanessa Raabe, Tamia Davis, Mary Olson

**Ohio State University AIDS Clinical Trials Unit – NIAID CoVPN, Columbus, OH:** Seuli Brill, Carlos Malvestutto, Susan Koletar, Taru Saigal, Mahdee Sobhanie, Vignesh Doraiswamy, Mahrous Abo Hassan, Jeremy Young

**<u>Orlando Immunology Center, Orlando, FL</u>:** Edwin DeJesus, Charlotte-Paige Rolle, Federico Hinestrosa, Dan Cruz, Terry Wilder, Jeffrey Garrett, Stephanie Skipper

**<u>Paradigm Clinical Research Institute, Torrance, CA</u>:** Ramprasad Dandillaya, Kartik Ananth

**<u>Penn Prevention – NIAID CoVPN Philadelphia, PA</u>:** Ian Frank, Helen Koenig, Eileen Donaghy, Debora Dunbar

**<u>PMG Research of McFarland Clinic, Ames, IA</u>:** Jennifer Killion, Rupal Amin, Shauna Basener, Timothy Lowry

**<u>PMG Research of Wilmington, Wilmington, NC</u>:** Kevin Cannon, Mesha Chadwick

**<u>Qway, Hialeah, FL</u>:** Oscar Galvez, Fausto Castillo

**<u>Regional One Health, Memphis, TN</u>:** John Jefferies, Sandy Arnold, Amber Thacker

**<u>Remington-Davis, Columbus, OH</u>:** Edward Cordasco, Brian Zeno, Heather Holmes, Heather Lee

**<u>Republican Clinical Hospital, Chisinau, Moldova</u>:** Natalia Gaibu, Victor Cojocaru, Aristia Seremet, Sergiu Iacob, Rodica Usatii, Nelea Ghicavii, Angela Coltuclu, Oxana Bujor

**<u>Rhode Island Hospital, Providence, RI</u>:** Eleftherios Mylonakis, Dimitrios Farmakiotis, Karen Tashima, Natasha Ryback

**<u>Ruane Clinical Research Group, Los Angeles, CA</u>:** Peter Ruane, Peter Wolfe, Kenny Trinidad

**<u>Rush University Medical Center, Chicago, IL</u>:** James Moy, Raj Shah, Bandi Sindhura, Beverly Sha

**<u>San Francisco Research Institute, San Francisco, CA</u>:** Mark Savant, Francis Hsiao, Edna Yee

**<u>Sarasota Memorial Hospital, Sarasota, FL</u>:** Manuel Gordillo, Rishi Bhattacharyya, Sudha Tallapragada, Annette Artau, Julie Larkin, Roberto Mercado, Michael Milam,

Natan Kraitman, Michael Lowry, Sarah Temple, Lenka Offner, Rabih Loutfi, Kirk Voelker, Marshall Frank, Ashley Grant

**SignatureCare Emergency Center – TC Jester, Houston, TX:** Alan Skolnick, Harold Minkowitz, David Leiman, Todd Price, Anatoli Krasko

**St. Hope Foundation, Bellaire, TX:** James Sims III, Manuel Vasquez, Kenneth Degazon, Katherine Asuncion

**Stanford University, Palo Alto, CA:** Jason Andrews, Aruna Subramanian, Upinder Singh, Yvonne Maldonado, Chaitan Khosla

**Tandem Clinical Research, Maitland, FL:** Esteban Olivera, Mayra Abreu

**Tandem Clinical Research, Marrero, LA:** Adil Fatakia, Marissa Miller, Kristen Clinton, Gary Reiss

**The Hope Clinic of Emory University – NIAID CoVPN, Decatur, GA:** Srilatha Edupuganti, Nadine Rouphael, Colleen Kelley, Varun Phadke, Cassie Grimsley Ackerley, Matthew Collins

**The Lundquist Institute, Torrance, CA:** Loren Miller, Timothy Hatlen

**The Ponce de Leon Center Clinical Research Site – NIAID CoVPN, Atlanta, GA:** Michael Chung, Colleen Kelley, Valeria Cantos Lucio, Carlos del Rio, Jeffrey Lennox, Sheetal Kandiah, Caitlin Moran, Anandi Sheth, Paulina Rebolledo, Nithin Gopalsamy, Divya Bhamidipati

**Triple O Research Institute PA, West Palm Beach, FL:** Olayemi Osiyemi, Jose A. Menajovsky-Chaves, Christina Campbell

**Tufts Medical Center, Boston, MA:** Andrew Strand, Andreas Klein, Debra Poutsiaka, Roberto Viau Colindres, Brian Chow, Cheleste Thorpe, Mary Hopkins, Jenn Chow, Rakhi Kohli, Jose Caro, Jeffrey Griffiths, Helen Boucher, Whitney Perry, Laura Kogelman, Yoav Golan, Tine Vindenes, Carlos Mendoza, Saba Mostafavi, Christhian Alejandro Cano Guerra, Paula Dabenigno, Bipin Malla

**Tulane University School of Medicine, New Orleans, LA:** Dahlene Fusco, Arnaud Drouin, Joshua Denson, Jerry Zifodya, Christine Bojanowski, Monika Dietrich, Stacy Drury

**University of Illinois at Chicago Project WISH – NIAID CoVPN, Chicago, IL:** Jesica Herrick, Richard Novak, Mahesh Patel

**Universal Medical and Research Center, LLC, Miami, FL:** Gerard Acloque, Agustin Martinez

**University at Buffalo, State University of New York, Buffalo, NY:** Sanjay Sethi, Brian Clemency, Rajesh Kunadharaju

**University of Arizona, Tucson, AZ:** Sairam Parthasarathy, Franz Rischard

**University of California Davis, Sacramento, CA:** Stuart Cohen, George Thompson, Hien Nguyen, Scott Crabtree

**University of Cincinnati, Cincinnati, OH:** Carl Fichtenbaum, Moises Huaman, Jaime Robertson

**University of Colorado School of Medicine, Aurora, CO:** Eric Simoes, Thomas Campbell, Poornima Ramanan, Hillary Dunlevy, Esther Benamu, Amiran Baduashvili, Martin Krsak, Steven Johnson, Lakshmi Chauhan, Erica Fredregil, Samantha Economos

**University of Miami - Miller School of Medicine, Miami, FL:** Gary Kleiner, Lilian Abbo, Bhavarth Shukla, Jennifer Gebbia, Maria Rodriguez

**University of Minnesota, Minneapolis, MN:** Anne-Marie Leuck, Mahsa Abassi, Matthew Pullen

**University of Mississippi, Jackson, MS:** Jose Lucar Lloveras, Leandro Mena, Luis Shimose Ciudad

**University of North Carolina, Chapel Hill, NC:** Jessica Lin, David Wohl, Christopher Hurt, William Fischer II, Kathleen Tompkins

**University of South Florida, Tampa, FL:** Kami Kim, Seetha Lakshmi, Charurut Somboonwit, Jason Wilson, Asa Oxner, Tiffany Vasey, Lucy Guerra

**University of Virginia, Charlottesville, VA:** William Petri, Katie Dykstra, Marianne Morrissey, Lejla Cesko, Jae Shin, Cirle Warren, Jennifer Sasson, Chelsea Marie, Debbie-Ann Shirley, Rebecca Carpenter, Gregory Madden, Danielle Donigan, Michelle Sutton, Cynthia Edwards, Elizabeth Brooks, Rebecca Wade, Samantha Simmons, Jennifer Pinnata

**University of Washington Medical Center, Seattle, WA:** Ruanne Barnabas, Shelly Karuna, Ann C. Collier, Julie McElrath, Janine Maenza, Adrienne Shapiro, Helen Stankiewicz-Karita, Helen Chu, Chandler Church

**University of Wisconsin, Madison, WI:** William Hartman, Joseph Connor, Robert Striker

**University of Texas Health Science Center, Tyler, TX:** Julie Philley, Megan Devine, Richard Yates, Steven Hickerson

**Vanderbilt Vaccine Clinical Research Site – NIAID CoVPN, Nashville, TN:** Spyros Kalams, Greg Wilson

**Virginia Commonwealth University School of Medicine, Richmond, VA:** Michael Donnenberg, Marjolein de Wit

**VitaLink Research, Gaffney, SC:** David Erb, Luis DeLaCruz, Supinder Channa

**Whitman-Walker Health – NIAID CoVPN, Washington, DC:** Sarah Henn, Megan Coleman, Lynsay MacLaren, Deborah Goldstein, Alice Eggleston, Carrington Koebele

**WR-ClinSearch, LLC, Chattanooga, TN:** Mark McKenzie, Teresa Deese

**WR-Mount Vernon Clinical Research, LLC, Sandy Springs, GA:** Benjamin Thomas, Laura Tsakiris, Stephen Blank, Ronald Mirenda

**Xera Med Research, Boca Raton, FL:** Anna Martin, Gargi Gharat, Candace Kokaram, Ket Wray, Clement Partap, Ulyana Arzamasova, Kristina Louissaint, Maria Fernandez

**Xera Med Research, Miami, FL:** Anna Martin, Ket Wray, Kristina Louissaint, Maria Fernandez, Gargi Gharat

**Regeneron Study Team**

Achint Chani, Adebiyi Adepoju, Adnan Mahmood, Aisha Mortagy, Ajla Dupljak, Alina Baum, Alison Brown, Amy Froment, Andrea Hooper, Andrea Margiotta, Andrew Bombardier, Anita Islam, Anne Smith, Arvinder Dhillon, Audra McMillian, Aurora Breazna, Ayesha Aslam, Barabara Carpentino, Bari Kowal, Barry Siliverstein, Benjamin Horel, Bo Zhu, Bret Musser, Brian Bush, Brian Head, Brian Snow, Bryan Zhu, Camille Debray, Careta Phillips, Carmella Simiele, Carol Lee, Carolyn Nienstedt, Caryn Trbovic, Casey (Kuo-Chen) Chan, Catherine Elliott, Chad Fish, Charlie Ni, Christa Polidori, Christine Enciso, Christopher Caira, Christopher Powell, Christos A. Kyratsous, Cliff Baum, Colin McDonald, Cynthia Leigh, Cynthia Pan, Dana Wolken, Danielle Manganello, David Liu, David Stein, David M. Weinreich, Dawlat Hassan, Daya Gulabani, Deborah Fix, Deborah Leonard, Deepshree Sarda, Denise Bonhomme, Denise Kennedy, Devin Darcy, Dhanalakshmi Barron, Diana Hughes, Diana Rofail, Dipinder Kaur, Divya Ramesh, Dona Bianco, Donna Cohen, Eduardo Forleo Neto, Edward Jean-Baptiste, Ehsan Bukhari, Eileen Doyle, Elizabeth Bucknam, Emily Labriola-Tomkins, Emily Nanna, Esther Huffman O'Keefe, Evelyn Gasparino, Evonne Fung, Flonza Isa, Fung-Yee To, Gary Herman, George D. Yancopoulos, Georgia Bellingham, Giane Sumner, Grainne Moggan, Grainne Power, Haixia Zeng, Hazel Mariveles, Heath Gonzalez, Helen Kang, Hibo Noor, Ian Minns, Ingeborg Heirman, Izabella Peszek, James Donohue, Jamie Rusconi, Janice Austin, Janie Parrino, Jeannie Yo, Jenna McDonnell, Jennifer D. Hamilton, Jessica Boarder, Jianguo Wei, Jingchun Yu, Joanne Malia, Joanne Tucciarone, Jodie Tyler-Gale, John D. Davis, John Strein, Jonathan Cohen, Jonathan Meyer, Jordan Ursino, Joseph Im, Joseph Tramaglini,

Joseph Wolken, Kaitlyn Potter, Kaitlyn Scacalossi, Kamala Naidu, Karen Browning, Karen Rutkowski, Karen Yau, Katherine Woloshin, Kelly Lewis-Amezcua, Kenneth Turner, Kimberly Dornheim, Kit Chiu, Kosalai Mohan, Kristina McGuire, Kristy Macci, Kurt Ringleben, Kusha Mohammadi, Kyle Foster, Latora Knighton, Leah Lipsich, Lindsay Darling, Lisa Boersma, Lisa Cowen, Lisa Hersh, Lisa Jackson, Lisa Purcell, Lisa Sherpinsky, Livia Lai, Lori Faria, Lori Geissler, Louise Boppert, Lyra Fiske, Marc Dickens, Marco Mancini, Maria Cynthia Leigh, Meagan O'Brien, Michael Batchelder, Michael Klinger, Michael Partridge, Michel Tarabocchia, Michelle Wong, Mivianisse Rodriguez, Moetaz Albizem, Muriel O'Byrne, Ned Braunstein, Neena Sarkar, Neil Stahl, Nicole Deitz, Nicole Memblatt, Nirav Shah, Nitin Kumar, Olga Herrera, Oluchi Adedoyin, Ori Yellin, Pamela Snodgrass, Patrick Floody, Paul D'Ambrosio, Paul (Xiaobang) Gao, Peijie Hou, Philippa Hearld, Qin Li, Rachel Kitchenoff, Rakiyya Ali, Ramya Iyer, Ravikanth Chava, Rinol Alaj, Rita Pedraza, Robert Hamlin, Romana Hosain, Ruchin Gorawala, Ryan White, Ryan Yu, Rylee Fogarty, S. Balachandra Dass, Sagarika Bollini, Samit Ganguly, Sandra DeCicco, Sanket Patel, Sarah Cassimaty, Selin Somersan-Karakaya, Shane McCarthy, Sharon Henkel, Shazia Ali, Shelley Geila Shapiro, Somang Kim, Soraya Nossoughi, Stephanie Bisulco, Steven Elkin, Steven Long, Sumathi Sivapalasingam, Susan Irvin, Susan Wilt, Tami Min, Tatiana Constant, Theresa Devins, Thomas DiCioccio, Thomas Norton, Travis Bernardo, Tzu-Chien Chuang, Victor (Jianguo) Wei, Vinh Nuce, Vishnu Battini, Wilson Caldwell, Xiaobang Gao, Xin Chen, Yanmei Tian, Yasmin Khan, Yuming Zhao, Yunji Kim

**NIAID/CoVPN Team**

Bonnie Dye (CoVPN), Christopher B. Hurt (CoVPN), Dale R. Burwen (NIAID), Dan H. Barouch (CoVPN), David Burns (NIAID), Elizabeth Brown (CoVPN), Katharine J. Bar (CoVPN), Mary Marovich (NIAID), Meredith Clement (CoVPN), Myron S. Cohen (CoVPN), Nirupama Sista (CoVPN), Ruanne V. Barnabas (CoVPN), Sheryl Zwerski (NIAID)

### Supplementary Methods

#### Stratification

Assignment of treatment group (1200 mg REGEN-COV or placebo) was stratified by the following prior to randomization:

1. Test results (positive, negative, or undetermined) of a local diagnostic assay for SARS-CoV-2 (e.g., molecular assay such as RT-PCR for SARS-CoV-2 or a SARS-CoV-2 antigen test) from appropriate samples (e.g., nasopharyngeal, oropharyngeal, nasal, or saliva)
2. Age:
    - ≥12 and <18 years
    - ≥18 and <50 years
    - ≥50 years

**Inclusion and Exclusion Criteria**

**Inclusion criteria**

A participant must meet the following criteria to be eligible for inclusion in the study:

1. Adult participants 18 years of age (irrespective of weight) and above at the signing of informed consent or adolescent participants ≥12 to <18 years of age, or pediatric participants <12 years of age at the signing of the assent (parent/guardian sign the informed consent)
2. Asymptomatic household contact with exposure to an individual with a diagnosis of SARS-CoV-2 infection (index case). To be included in the study, participants must be randomized within 96 hours of collection of the index cases' positive SARS-COV-2 diagnostic test sample
3. Participant anticipates living in the same household with the index case until study day 29
4. Is judged by the investigator to be in good health based on medical history and physical examination at screening/baseline, including participants who are healthy or have a chronic, stable medical condition
5. Willing and able to comply with study visits and study-related procedures/assessments
6. Provide informed consent signed by study participant or legally acceptable representative

**Exclusion criteria**

A participant who meets any of the following criteria will be excluded from the study:

1. Participant-reported history of prior positive SARS-CoV-2 RT-PCR test or positive SARS CoV-2 serology test at any time before the screening
2. Participant has lived with individuals who have had previous SARS-CoV-2 infection or currently lives with individuals who have SARS-CoV-2 infection, with the exception of the index case(s), the first individual(s) known to be infected in the household
3. Active respiratory or non-respiratory symptoms consistent with COVID-19
4. History of respiratory illness with sign/symptoms of SARS-CoV-2 infection, in the opinion of the investigator, within the prior 6 months to screening
5. Nursing home resident
6. Any physical examination findings, and/or history of any illness, concomitant medications or recent live vaccines that, in the opinion of the study investigator, might confound the results of the study or pose an additional risk to the participant by their participation in the study
7. Current hospitalization or was hospitalized (i.e., >24 hours) for any reason within 30 days of the screening visit
8. Has a history of significant multiple and/or severe allergies (e.g., latex gloves), or has had an anaphylactic reaction to prescription or non-prescription drugs or food. This is to avoid possible confounding of the safety analysis and not due to any presumed increased risk of these individuals to a reaction to the investigational product

9. Treatment with another investigational agent in the last 30 days or within five half-lives of the investigational drug, whichever is longer, prior to the screening visit
10. Received an investigational or approved SARS-CoV-2 vaccine
11. Received investigational or approved passive antibodies for SARS-CoV-2 infection prophylaxis (e.g., convalescent plasma or sera, monoclonal antibodies, hyperimmune globulin)
12. Use of hydroxychloroquine/chloroquine for prophylaxis/treatment of SARS-CoV-2 or anti-SARS-viral agents,* e.g., remdesivir, within 60 days of screening

    *Hydroxychloroquine/chloroquine for other uses, e.g.,, for use in autoimmune diseases, is allowed
13. Member of the clinical site study team and/or immediate family
14. Exclusion criterion #14 excluding sexually active men who are unwilling to use the following forms of medically acceptable birth control during the study drug follow-up period and for 8 months after single dose of study drug was removed since enrollment was expanded to include all women in protocol amendment 4
15. Exclusion criterion #15 excluding pregnant or breastfeeding women was removed since enrollment was expanded to all women in protocol amendment 4
16. Exclusion criterion #16 excluding women of childbearing potential (WOCBP)* and girls at or beyond menarche (≥12 to <18 years of age) who were unwilling to practice highly effective contraception prior to the initial dose/start of the first treatment, during the study, and for at least 8 months after the last dose was removed since enrollment was expanded to all women in protocol amendment 4

*WOCBP are defined as women who are fertile following menarche until becoming postmenopausal, unless permanently sterile. Permanent sterilization methods include hysterectomy, bilateral salpingectomy, and bilateral oophorectomy

**Strict-Term, Broad-Term, and CDC Definitions of COVID-19 Signs and Symptoms**

**Strict-Term**

Fever (≥38°C) PLUS ≥1 respiratory symptom (sore throat, cough, or shortness of breath)

*OR*

Two respiratory symptoms (sore throat, cough, or shortness of breath)

*OR*

One respiratory symptom (sore throat, cough, or shortness of breath) PLUS ≥2 non-respiratory symptoms (chills, nausea, vomiting, diarrhea, headache, conjunctivitis, myalgia, arthralgia, loss of taste or smell, fatigue, or general malaise)

**Broad-Term**

Fever ≥38°C

The signs and symptoms below:

1. Feverish
2. Sore throat
3. Cough
4. Shortness of breath/difficulty breathing (*nasal flaring**)
5. Chills

6. Nausea
7. Vomiting
8. Diarrhea
9. Headache
10. Red or watery eyes (*conjunctivitis*)
11. Body aches such as muscle pain or joint pain (*myalgia, arthralgia*)
12. Loss of taste/smell
13. Fatigue (*fatigue or general malaise or lethargy**)
14. Loss of appetite or poor eating/feeding
15. Confusion
16. Dizziness
17. Pressure/tightness in chest
18. Chest pain
19. Stomach ache (*abdominal pain**)
20. Rash
21. Sneezing
22. Runny nose
23. Sputum/phlegm
24. Other

*Signs and symptoms observed in pediatric participants

**CDC Definition**

At least two of the following symptoms: fever (measured or subjective), chills, rigors, myalgia, headache, sore throat, nausea or vomiting, diarrhea, fatigue, congestion, or runny nose

OR

Any one of the following symptoms: cough, shortness of breath, difficulty breathing, new olfactory disorder, or new taste disorder

OR

Severe respiratory illness with at least one of the following, clinical or radiographic evidence of pneumonia, or acute respiratory distress syndrome.

#### Trial Oversight

Regeneron designed the trial in collaboration with CoVPN and NIAID, and gathered the data with the trial investigators. Regeneron analyzed the data. The investigators, site personnel, CoVPN/NIAID, and Regeneron were blinded to treatment-group assignments. A Data and Safety Monitoring Board convened by the National Institutes of Health evaluated safety data to make recommendations for trial modification and/or termination.

The trial was conducted in accordance with the principles of the Declaration of Helsinki, the International Council for Harmonisation Good Clinical Practice guidelines, and all applicable regulatory requirements. The central or local institutional review board or ethics committee at each study center oversaw trial conduct and documentation. All participants provided written informed consent before participating in the trial.

#### Additional Statistical Methods

##### Analyses for Key Secondary Efficacy Endpoints

For the key secondary efficacy endpoints, the binary endpoints of proportion of participants were analyzed using the logistic regression similar to the primary efficacy endpoint. The continuous endpoints for duration of events were analyzed by Van Elteren test stratified by region (US vs. ex-US) and age (≥12 to <50 years vs. ≥50 years). The endpoint of proportion of participants with index case linkage to REGEN-COV treatment in study COV-2067 was analyzed by Fisher exact test.

##### Missing Data Imputation

Participants with missing central lab RT-qPCR test results were considered as symptomatic infections if any symptoms occurred within 14 days of a positive SARS-CoV-2 test from a local lab.

For viral load endpoints, if nasopharyngeal swab viral load data is missing for a visit, it is not imputed regardless of symptoms. Only non-missing available nasopharyngeal swab viral load data are used for the analysis for viral load endpoints. Only participants with at least one post-baseline viral load data in nasopharyngeal swab samples are included in the analysis.

**Pharmacokinetic Analysis Methods**

**Bioanalytical Methods**

The human serum concentrations of REGN10933 (casirivimab) and REGN10987 (imdevimab) were measured using validated immunoassays which employ streptavidin microplates from Meso Scale Discovery (MSD, Gaithersburg, MD, USA). The methods utilized two anti-idiotypic monoclonal antibodies, each specific for either REGN10933 or REGN10987, as the capture antibodies. Captured REGN10933 and REGN10987 were detected using two different, non-competing anti-idiotypic monoclonal antibodies, each also specific for either REGN10933 or REGN10987. The bioanalytical methods specifically quantitated the levels of each anti-SARS-CoV-2 spike monoclonal antibody separately, with no interference from the other antibody. The assay has a lower limit of quantification of 0.156 mg/L for each analyte in the undiluted serum sample.

**Pharmacokinetic Methods**

The pharmacokinetic (PK) analysis population included all participants who received any study drug (safety population) and who had at least one non-missing result following the first dose of study drug. Participants were analyzed based on actual treatment received. For the sentinel group of Part A, 12 participants were included in the PK analysis set. For the safety group of Part A, 89 participants were included in the PK analysis set. Overall, the PK analysis set is comprised of 101 participants.

Blood samples for measurement of casirivimab and imdevimab concentrations in serum were collected from the first 30 participants randomized to 1200 mg SC or placebo (sentinel group, subset 1) at predose and on Days 2, 4, 8, 15, 22, 29, 57, 85,

113, 141, 169, 197, and 225. Blood samples for drug concentration also were collected in the 31st to 400th participants randomized to 1200 mg or placebo (safety group, subset 2) at predose and on Days 29, 57, 113, 169, and 225.

From the above drug concentration sampling schedule, PK parameters were determined by non-compartmental methods (Gibaldi 1982) using Phoenix WinNonlin (Certara Corporation). Area under the curve in serum from day 0 to 28 ($AUC_{0\text{-}28}$) was determined using the log-linear trapezoidal rule and actual sample collection times. Area under the curve from time zero extrapolated to infinite time ($AUC_{inf}$) was determined as area under the curve through the last measurable concentration ($C_{last}$) + $C_{last}/\lambda_z$, where $\lambda_z$ is the slope of the terminal phase of the concentration–time curve. $C_{max}$ was determined as the maximum observed concentration in serum and $t_{max}$ was the time of maximal concentration in serum. The observed half-life ($t_{1/2}$) was estimated using actual times.

### Supplementary Results

#### Initial Descriptive Assessment

As this phase 3 trial was conducted without any prior phase 2 prevention data, an initial descriptive assessment (without formal testing) of the first 554 participants in Part A was conducted to verify the assumptions made during study planning, estimate the sample size, and inform the phase 3 analysis plans (as detailed in the statistical analysis plan). In 409 out of 554 individuals without current or prior infection (RT-qPCR-negative/seronegative), REGEN-COV reduced symptomatic infection by 100% (0% [0/186] vs. 3.6% [8/223], REGEN-COV vs. placebo, respectively) and overall (symptomatic and asymptomatic) infection by 47.9% (5.4% [10/186] vs. 10.3% [23/223], REGEN-COV vs. placebo, respectively). REGEN-COV also reduced the total duration of transmissibility (9 weeks vs. 44 weeks of infection, and 0 vs. 22 weeks of high viral load [>$10^4$ copies/mL] in the REGEN-COV and placebo groups, respectively).

**Impact of Treatment of Index Cases on Household Contacts**

We sought to understand whether treatment of an index case with REGEN-COV would impact rates of infection, symptomatic or asymptomatic, in household contacts in this study. No apparent differences in infection rates were observed. Among household contacts receiving placebo in this study, 23/116 (19.8%) lived with an index case (first known household member with SARS-CoV-2 infection) who received REGEN-COV in study COV-2067 while 10/51 (19.6%) lived with an index case who received placebo (participants were randomized 2:1 REGEN-COV to placebo). Among household contacts receiving REGEN-COV in this study, 6/113 (5.3%) and 3/60 (5.0%) resided with an index case who received REGEN-COV or placebo, respectively, in study COV-2067.

### Supplementary Figures

#### Figure S1. Schematic Overview of the Study Design

**Eligible participants:**

- Age criterion: adults (≥18 years of age), adolescents (≥12 and <18 years of age), and pediatrics (<12 years of age)*
- Asymptomatic household contact with exposure to an individual with a diagnosis of SARS-CoV-2 infection (index case)
- Randomized within 96 hours of collection of the index case's positive test sample
- Baseline RT-qPCR test to determine if in Part A (RT-qPCR negative) or Part B (RT-qPCR positive) for data analysis

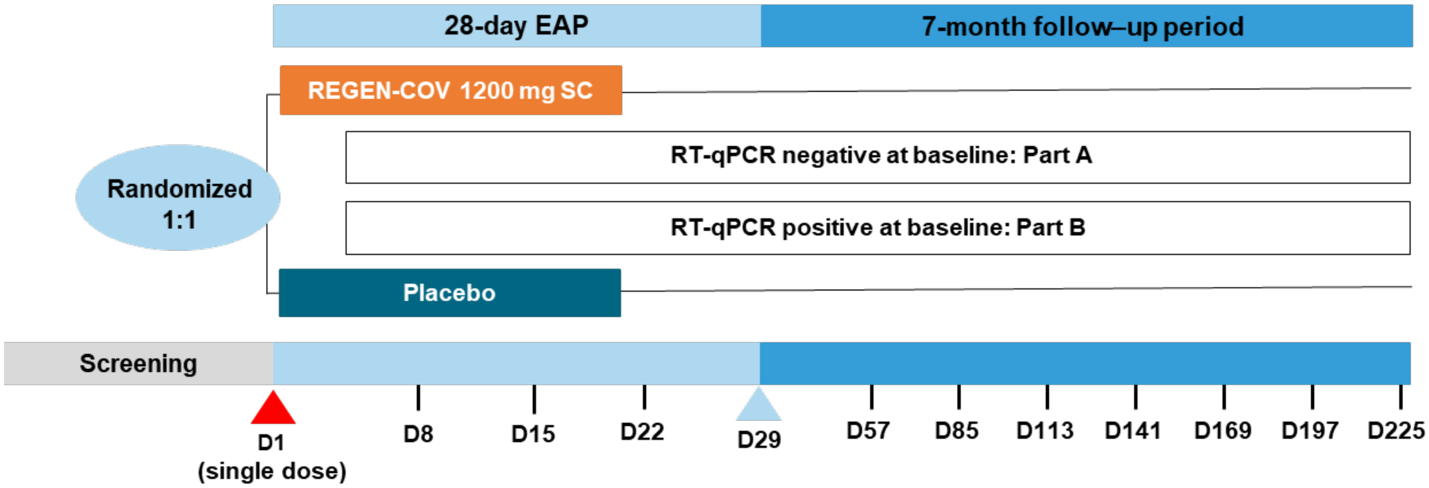

*Pediatric participants are not included in the presented analysis. Covid-19 denotes coronavirus disease 2019, D day, EAP efficacy assessment period, RT-qPCR quantitative reverse transcription polymerase chain reaction, and SC subcutaneous.

#### Figure S2. Flow Diagram for the Analysis Population

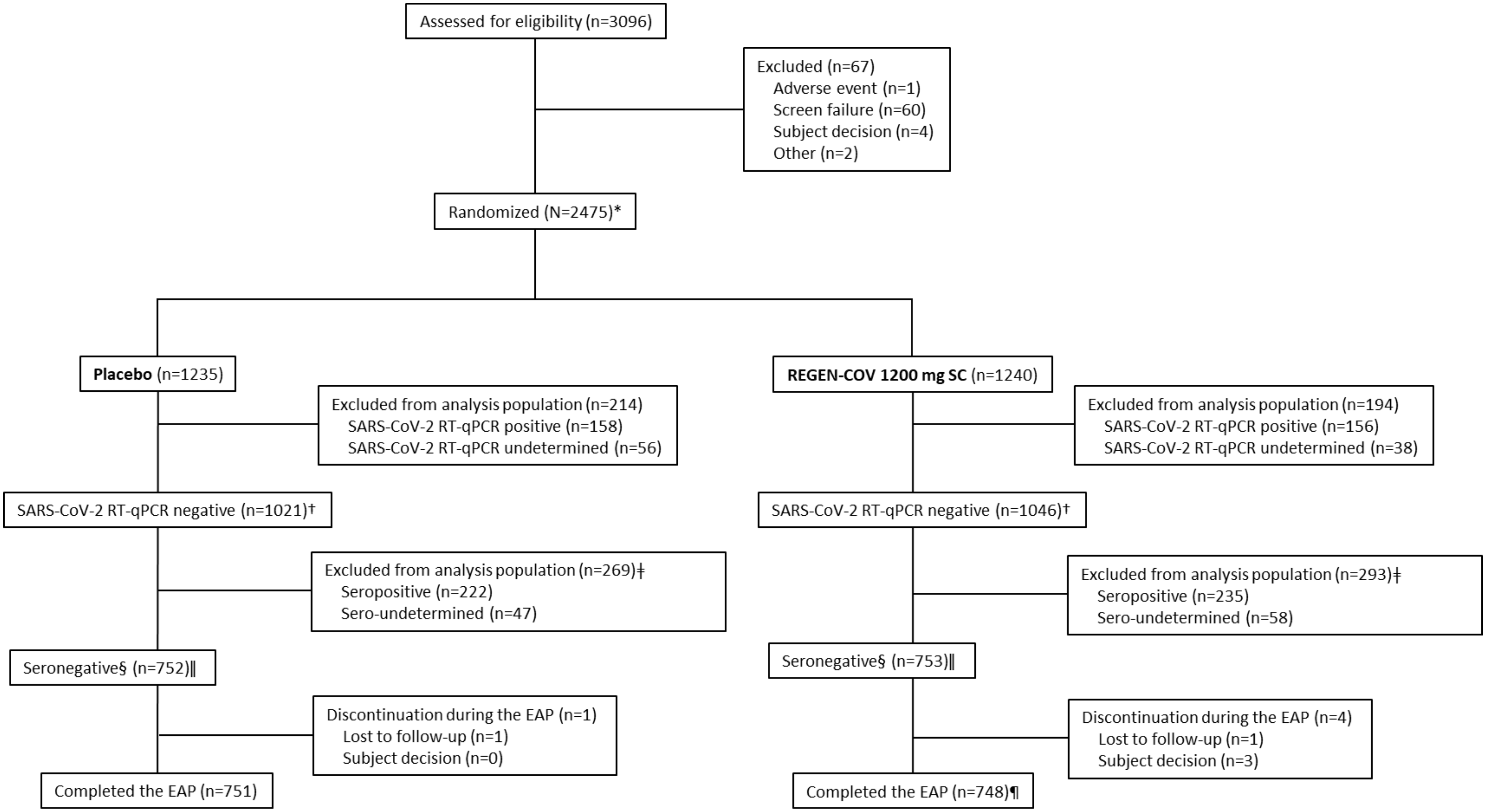

*Excludes the 554 participants from the administrative analysis for efficacy analyses; these particpants were included in the safety analysis population. EAP denotes efficacy assessment period, mFAS modified full analysis set, RT-qPCR quantitative reverse transcription polymerase chain reaction, and SC subcutaneous.

†This is the mFAS-A population.
‡Seropositive means one or more positive results from an anti-SARS-CoV-2 antibody test; sero-undetermined means there was a borderline positive anti-SARS-CoV-2 antibody test.
§Seronegative means no evidence of prior infection (no evidence of anti-SARS-CoV-2 antibodies).
‖This is the seronegative mFAS-A (primary efficacy analysis) population.
¶One participant was randomized but not treated and therefore did not complete the EAP but was included in the seronegative mFAS-A population.

**Figure S3. Heatmap of High SARS-CoV-2 Viral Load Infection (>$10^4$ Copies/mL) Over Time**

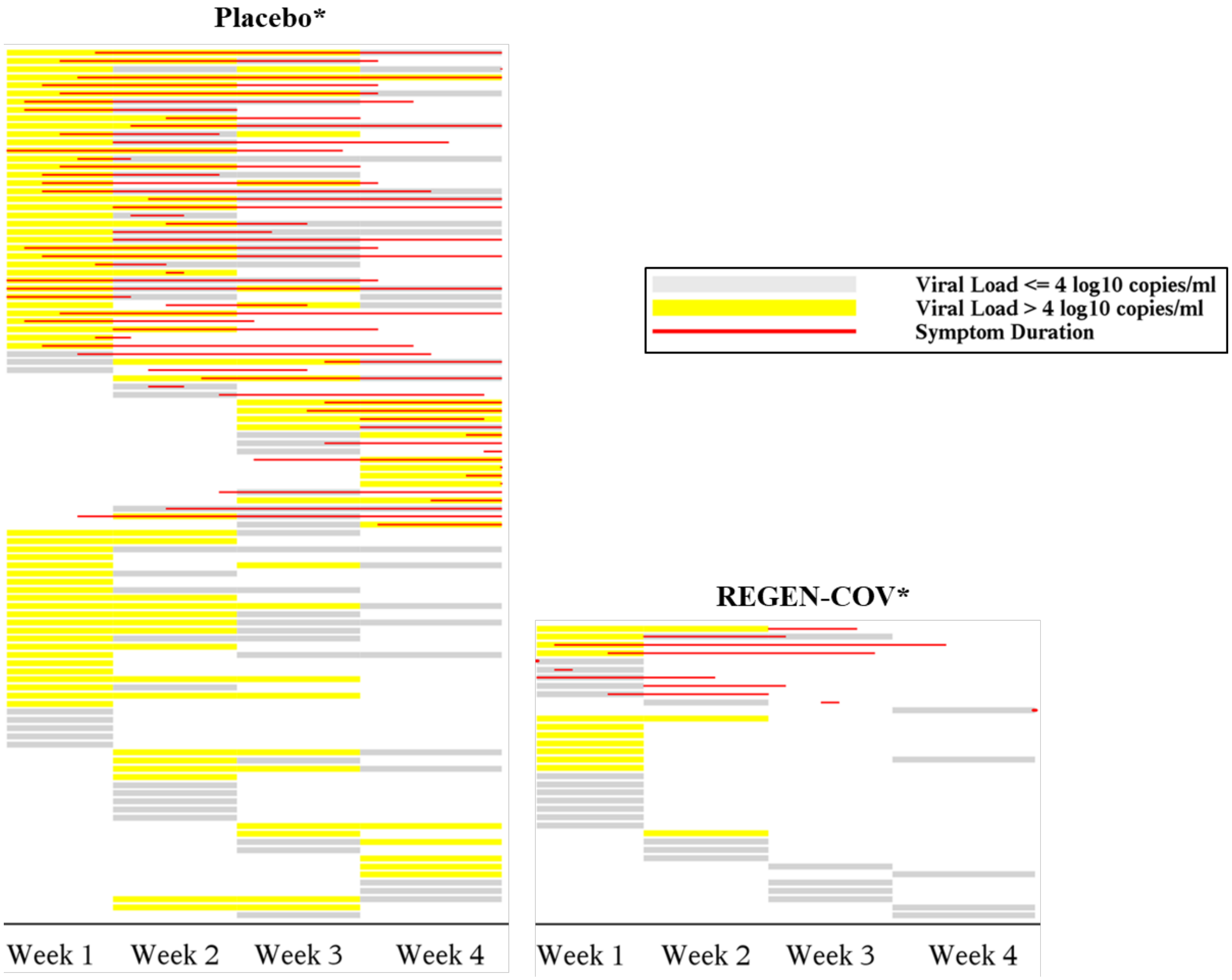

*Each row represents an individual participant with SARS-CoV-2 infection.

**Figure S4. Mean (±SD) Concentrations of Casirivimab (REGN10933) and Imdevimab (REGN10987) in Serum Over Time in Adults with Household Contact Exposure to Individuals with SARS-CoV-2 Infection (Part A Sentinel Group)**

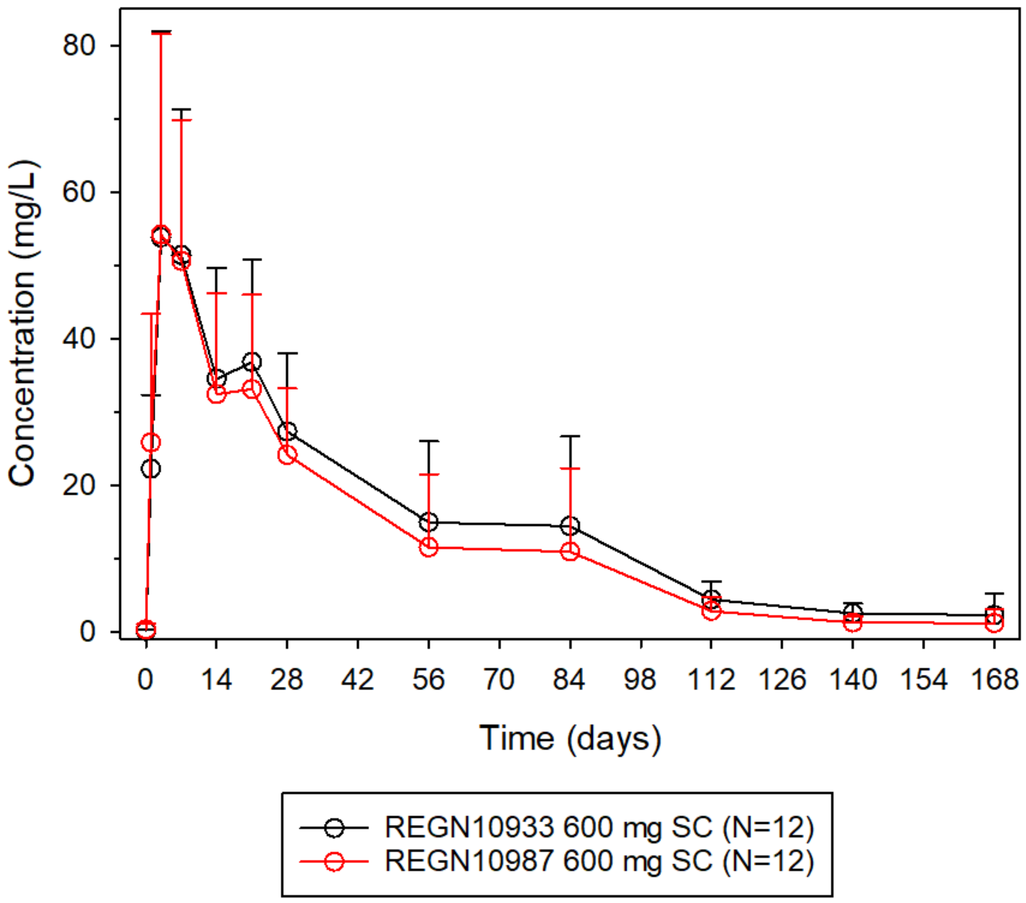

SC denotes subcutaneous, and SD standard deviation.

#### Supplementary Tables

##### Table S1. Hierarchy Testing Sequence of Key Secondary Efficacy Endpoints

| Sequence | Key Secondary Efficacy Endpoint |
|---|---|
| 1 | Proportion of participants with viral load >4 $log_{10}$ copies/mL in nasopharyngeal swab samples during the EAP |
| 2 | Number of weeks of symptomatic RT-qPCR confirmed SARS-CoV-2 infection (broad-term) during the EAP |
| 3 | Number of weeks of high-viral load >4 $log_{10}$ copies/mL in nasopharyngeal swab samples during the EAP |
| 4 | Number of weeks of RT-qPCR confirmed SARS-CoV-2 infection (regardless of symptoms) during the EAP |
| 5 | Proportion of participants who have a RT-qPCR confirmed SARS-CoV-2 infection (regardless of symptoms) during the EAP |
| 6 | Proportion of participants in placebo group with a RT-qPCR confirmed SARS-CoV-2 infection during the EAP with an index case participating in study COV-2067 (comparison of those whose index cases receive REGEN-COV versus placebo in study COV-2067) |

EAP denotes efficacy assessment period, and RT-qPCR quantitative reverse transcription polymerase chain reaction.

#### Table S2. Demographics and Baseline Characteristics (Seropositive)

| | Placebo (N=222) | REGEN-COV 1200 mg SC (N=235) | Total (N=457) |
|---|---|---|---|
| Age – years | | | |
| Mean (range) | 41.5 (12–80) | 40.6 (12–77) | 41.0 (12–80) |
| ≥50 – no. (%) | 72 (32.4) | 72 (30.6) | 144 (31.5) |
| Male sex – no. (%) | 108 (48.6) | 114 (48.5) | 222 (48.6) |
| Race – no. (%) | | | |
| White | 189 (85.1) | 179 (76.2) | 368 (80.5) |
| Black or African American | 27 (12.2) | 45 (19.1) | 72 (15.8) |
| Asian | 2 (0.9) | 7 (3.0) | 9 (2.0) |
| Native Hawaiian or Pacific Islander | 0 | 1 (0.4) | 1 (0.2) |
| Other | 4 (1.8) | 3 (1.3) | 7 (1.5) |
| Ethnicity — no. (%) | | | |
| Hispanic or Latino | 150 (67.6) | 161 (68.5) | 311 (68.1) |
| Not Hispanic or Latino | 72 (32.4) | 74 (31.5) | 146 (31.9) |
| Other | 0 | 0 | 0 |
| Mean weight — kg | 82.8±19.58 | 80.8±18.95 | 81.8±19.26 |
| Body-mass index† | | | |
| Mean | 29.4±6.18 | 28.7±6.29 | 29.0±6.24 |
| >30 – no. (%) | 85 (38.3) | 88 (37.4) | 173 (37.9) |
| Participants with any high-risk factor for Covid-19 – no. (%) | 63 (28.4) | 61 (26.0) | 124 (27.1) |
| ≥65 years of age | 18 (8.1) | 14 (6.0) | 32 (7.0) |
| Body-mass index† ≥35 kg/m$^2$ | 38 (17.1) | 35 (14.9) | 73 (16.0) |
| Chronic kidney disease | 2 (0.9) | 1 (0.4) | 3 (0.7) |
| Diabetes | 12 (5.4) | 13 (5.5) | 25 (5.5) |
| Immunosuppressive disease | 0 | 2 (0.9) | 2 (0.4) |
| Receiving immunosuppressive treatment | 0 | 1 (0.4) | 1 (0.2) |
| ≥55 years of age with CVD, hypertension, or COPD | 20 (9.0) | 19 (8.1) | 39 (8.5) |
| Total no. of households | 213 | 228 | 413 |
| Number of households by size – no. (%)‡ | | | |
| 1 | 180 (84.5) | 195 (85.5) | 375 (90.8) |
| 2 | 27 (12.7) | 27 (11.8) | 32 (7.7) |
| 3 | 6 (2.8) | 6 (2.6) | 6 (1.5) |
| 4 | 0 | 0 | 0 |
| >4 | 0 | 0 | 0 |

| | | | |
|---|---|---|---|
| Participants with an index case participating in study COV-2067– no. (%) | 38 (17.1) | 32 (13.6) | 70 (15.3) |

*Plus-minus values are means ±SD. BMI denotes body mass index, COPD chronic obstructive pulmonary disease, CVD cardiovascular disease, SC subcutaneous, and SD standard deviation.
†The body-mass index is the weight in kilograms divided by the square of the height in meters.
‡Household size is calculated by counting the seronegative study participants in Part A. Percentages are based on the number of households as the denominator, instead of the number of participants.

**Table S3. Proportion of Participants Who Have a Symptomatic RT-qPCR-Confirmed SARS-CoV-2 Infection (Broad-Term) by Week**

| | Placebo (N=752) | REGEN-COV 1200 mg SC (N=753) | Relative Risk Difference |
|---|---|---|---|
| Week 1 | 32 (4.3%) | 9 (1.2%) | 71.9% |
| Week 2 | 13 (1.7%) | 0 (0%) | |
| Week 3 | 7 (0.9%) | 1 (0.1%) | 92.6% |
| Week 4 | 7 (0.9%) | 1 (0.1%) | |

RT-qPCR denotes quantitative reverse transcription polymerase chain reaction, and SC subcutaneous.

**Table S4. Proportion of Participants Who Have a Symptomatic RT-qPCR-Confirmed SARS-CoV-2 Infection by Definition**

| | Placebo (N=752) | REGEN-COV 1200 mg SC (N=753) |
|---|---|---|
| Broad-term symptomatic infection* | | |
| n/N (%) | 59/752 (7.8) | 11/753 (1.5) |
| Relative risk reduction | - | 81.4% |
| Odds ratio (95% CI)† | - | 0.17 (0.09, 0.33) |
| P-value† | - | <0.0001 |
| CDC definition of symptomatic infection | | |
| n/N (%) | 46/752 (6.1) | 6/753 (0.8) |
| Relative risk reduction | - | 87.0% |
| Odds ratio (95% CI)† | - | 0.12 (0.05, 0.29) |
| P-value† | - | <0.0001 |
| Strict-term symptomatic infection | | |
| n/N (%) | 22/752 (2.9) | 2/753 (0.3) |
| Relative risk reduction | - | 90.9% |
| Odds ratio (95% CI)† | - | 0.09 (0.02, 0.37) |
| P-value† | - | 0.0010 |

*Primary end point. CDC denotes Centers for Disease Control and Prevention, CI confidence interval, RT-qPCR quantitative reverse transcription polymerase chain reaction, and SC subcutaneous.
†Based on logistic regression model adjusted by region (US vs. ex-US) and age group (12 to <50 vs. ≥50 years).

**Table S5. Proportion of Participants Who Have a Symptomatic RT-qPCR-Confirmed SARS-CoV-2 Infection (Broad-Term) by Baseline Serology Status**

| | Placebo | REGEN-COV 1200 mg SC |
|---|---|---|
| Seronegative* | | |
| n/N (%) | 59/752 (7.8) | 11/753 (1.5) |
| Relative risk reduction | - | 81.4% |
| Odds ratio (95% CI)† | - | 0.17 (0.09, 0.33) |
| P-value | - | <0.0001 |
| Seropositive | | |
| n/N (%) | 5/222 (2.3) | 1/235 (0.4) |
| Relative risk reduction | - | 81.1% |
| Odds ratio (95% CI)† | - | 0.19 (0.02, 1.68) |
| P-value | - | 0.1369 |
| Seronegative, seropositive, and sero–undetermined | | |
| n/N (%) | 66/1021 (6.5) | 12/1046 (1.1) |
| Relative risk reduction | - | 82.3% |
| Odds ratio (95% CI)† | - | 0.17 (0.09, 0.31) |
| P-value | - | <0.0001 |

*Primary end point. CI denotes confidence interval, RT-qPCR quantitative reverse transcription polymerase chain reaction, and SC subcutaneous.
†Based on the logistic regression model adjusted by region (US vs. ex-US) and age group (12 to <50 vs. ≥50 years).

#### Table S6. Proportion of Participants with SARS-CoV-2 Infection by Viral Load Category

| | Placebo (N=749) | REGEN-COV 1200 mg SC (N=745) |
|---|---|---|
| qPCR >$10^3$ copies/mL | | |
| n/N (%) | 92/749 (12.3) | 28/745 (3.8) |
| Relative risk reduction | - | 69.4% |
| Odds ratio (95% CI)* | - | 0.28 (0.18, 0.43) |
| qPCR >$10^4$ copies/mL | | |
| n/N (%) | 85/749 (11.3) | 12/745 (1.6) |
| Relative risk reduction | - | 85.8% |
| Odds ratio (95% CI)* | - | 0.13 (0.07, 0.24) |
| qPCR >$10^5$ copies/mL | | |
| n/N (%) | 78/749 (10.4) | 5/745 (0.7) |
| Relative risk reduction | - | 93.6% |
| Odds ratio (95% CI)* | - | 0.06 (0.02, 0.14) |

*Based on logistic regression model adjusted by region (US vs. ex-US) and age group (12 to <50 vs. ≥50 years). qPCR denotes quantitative polymerase chain reaction, and SC subcutaneous.

#### Table S7. Summary of Secondary Viral Load Endpoints Among Participants With a Positive RT-qPCR (Seronegative)

| Endpoint | Placebo (N=107) | REGEN-COV 1200 mg SC (N=36) |
|---|---|---|
| Time-weighted average of viral load ($log_{10}$ copies/mL) from the first positive SARS-CoV-2 RT-qPCR until the second weekly visit after the first positive test* | | |
| n | 98 | 33 |
| Mean (SD) | 3.79 (1.88) | 1.34 (0.93) |
| LS mean difference (SE) vs. placebo [95% CI]† | - | -2.48 (0.34) [-3.16 to -1.81] |
| Time-weighted average of viral load ($log_{10}$ copies/mL) from the first positive SARS-CoV-2 RT-qPCR until the third weekly visit after the first positive test* | | |
| n | 98 | 33 |
| Mean (SD) | 3.07 (1.65) | 0.99 (0.63) |
| LS mean difference (SE) vs. placebo [95% CI]† | - | -2.12 (0.30) [-2.71 to -1.54] |
| Maximum SARS-CoV-2 RT-qPCR viral load ($log_{10}$ copies/mL) among individuals with ≥1 positive RT-qPCR* | | |
| n | 107 | 36 |
| Mean (SD) | 6.41 (2.20) | 3.99 (1.26) |
| LS mean difference (SE) vs. placebo [95% CI]† | - | -2.43 (0.39) [-3.20 to -1.66] |
| SARS-CoV-2 RT-qPCR viral load ($log_{10}$ copies/mL) corresponding to the onset of first positive RT-qPCR* | | |
| n | 104 | 36 |
| Mean (SD) | 6.39 (2.149) | 3.96 (1.28) |
| LS mean difference (SE) vs. placebo [95% CI]† | - | -2.44 (0.38) [-3.19 to -1.69] |
| Area under the curve in viral load ($log_{10}$ copies/mL·day) from the first positive SARS-CoV-2 RT-qPCR until the first confirmed negative test*‡ | | |
| n | 81 | 33 |
| Mean (SD) | 65.98 (47.69) | 19.70 (14.83) |
| LS mean difference (SE) vs. placebo [95% CI]† | - | -47.68 (8.65) [-64.82 to -30.53] |

*In nasopharyngeal swab samples, that has an onset/during the efficacy assessment period. LS denotes least-squares, RT-qPCR quantitative reverse transcription polymerase chain reaction, SC subcutaneous, SD standard deviation, and SE standard error.

†Least-squares mean and standard error taken from the analysis of variance method with the fixed categorical effects of treatment group, age group (≥12 to <50 vs. ≤50 years), and region (US vs. ex-US).

‡For these endpoints, data collection on viral load is ongoing in the follow-up period.

#### Table S8. Overview of Treatment-Emergent Adverse Events

| | Placebo (N=1306) | | REGEN-COV 1200 mg SC (N=1311) | |
|---|---|---|---|---|
| no. (%) | Overall | Non-Covid-19 | Overall | Non-Covid-19 |
| Number of TEAEs | 709 | 481 | 556 | 483 |
| Number of grade ≥3 TEAEs | 25 | 19 | 21 | 21 |
| Number of serious TEAEs | 17 | 11 | 14 | 14 |
| Number of AESIs | 0 | 0 | 0 | 0 |
| Number of TEAEs resulting in study drug being withdrawn | 0 | 0 | 0 | 0 |
| Number of TEAEs resulting in death | 2 | 2 | 2 | 2 |
| Participants with at least one TEAE | 379 (29.0) | 215 (16.5) | 265 (20.2) | 210 (16.0) |
| Participants with at least one grade ≥3 TEAE | 22 (1.7) | 17 (1.3) | 11 (0.8) | 11 (0.8) |
| Participants with at least one serious TEAE | 15 (1.1) | 10 (0.8) | 10 (0.8) | 10 (0.8) |
| Participants with at least one AESI | 0 | 0 | 0 | 0 |
| Participants with at least one TEAE resulting in study drug being withdrawn | 0 | 0 | 0 | 0 |
| Participants with at least one TEAE resulting in death | 2 (0.2) | 2 (0.2) | 2 (0.2) | 2 (0.2) |

AESI denotes adverse event of special interest, SC subcutaneous, and TEAE treatment-emergent adverse event.

#### Table S9. Serious Adverse Events

| System Organ Class<br>Preferred Term – no. of participants (%) | Placebo<br>(N=1306) | REGEN-COV 1200 mg SC<br>(N=1311) |
|---|---|---|
| Participants with at least one serious TEAE | 15 (1.1) | 10 (0.8) |
| Infections and infestations | 9 (0.7) | 4 (0.3) |
| Gastroenteritis | 0 | 1 (<0.1) |
| Pneumonia | 1 (<0.1) | 1 (<0.1) |
| Sepsis | 0 | 1 (<0.1) |
| Soft tissue infection | 0 | 1 (<0.1) |
| Appendicitis | 1 (<0.1) | 0 |
| Covid-19 | 4 (0.3) | 0 |
| Covid-19 pneumonia | 2 (0.2) | 0 |
| Scrotal abscess | 1 (<0.1) | 0 |
| Urinary tract infection | 1 (<0.1) | 0 |
| Cardiac disorders | 1 (<0.1) | 1 (<0.1) |
| Acute myocardial infarction | 0 | 1 (<0.1) |
| Cardiac failure congestive | 0 | 1 (<0.1) |
| Cardiac arrest | 1 (<0.1) | 0 |
| Gastrointestinal disorders | 1 (<0.1) | 1 (<0.1) |
| Abdominal pain upper | 0 | 1 (<0.1) |
| Abdominal pain | 1 (<0.1) | 0 |
| General disorders and administration-site conditions | 0 | 1 (<0.1) |
| Sudden death | 0 | 1 (<0.1) |
| Hepatobiliary disorders | 0 | 1 (<0.1) |
| Cholecystitis acute | 0 | 1 (<0.1) |
| Injury, poisoning, and procedural complications | 1 (<0.1) | 1 (<0.1) |
| Ankle fracture | 0 | 1 (<0.1) |
| Foot fracture | 0 | 1 (<0.1) |
| Tibia fracture | 0 | 1 (<0.1) |
| Gunshot wound | 1 (<0.1) | 0 |
| Neoplasms benign, malignant and unspecified (incl. cysts and polyps) | 1 (<0.1) | 1 (<0.1) |
| Cervix carcinoma recurrent | 0 | 1 (<0.1) |
| Breast cancer | 1 (<0.1) | 0 |
| Respiratory, thoracic, and mediastinal disorders | 0 | 1 (<0.1) |
| Respiratory failure | 0 | 1 (<0.1) |
| Psychiatric disorders | 2 (0.2) | 0 |
| Mania | 1 (<0.1) | 0 |
| Suicidal ideation | 1 (<0.1) | 0 |

| | | |
|---|---|---|
| Vascular disorders | 1 (<0.1) | 0 |
| Essential hypertension | 1 (<0.1) | 0 |

SC denotes subcutaneous, and TEAE treatment-emergent adverse event.

#### Table S10. Deaths

| System Organ Class<br>Preferred Term, no. of participants (%) | Placebo<br>(N=1306) | REGEN-COV 1200 mg SC<br>(N=1311) |
|---|---|---|
| Participants with at least one TEAE leading to death | 2 (0.2) | 2 (0.2) |
| Cardiac disorders | 1 (<0.1) | 1 (<0.1) |
| Cardiac failure congestive | 0 | 1 (<0.1) |
| Cardiac arrest | 1 (<0.1) | 0 |
| General disorders and administration-site conditions | 0 | 1 (<0.1) |
| Sudden death | 0 | 1 (<0.1) |
| Injury, poisoning, and procedural complications | 1 (<0.1) | 0 |
| Gunshot wound | 1 (<0.1) | 0 |

SC denotes subcutaneous, and TEAE treatment-emergent adverse event.

**Table S11. Summary of PK Parameters for Casirivimab and Imdevimab After a Single 1200 mg SC Dose of REGEN-COV* in Part A Participants**

| PK Parameter† | Casirivimab (REGN10933) | Imdevimab (REGN10987) |
|---|---|---|
| $C_{max}$ (mg/L) | 58.5 (24.5) [11] | 55.2 (25.0) [11] |
| $t_{max}$ (day) ‡ | 8.0 (4.0, 87.0) [11] | 7.0 (4.0, 15.0) [11] |
| $AUC_{0-28}$ (mg●day/L) | 1099 (406) [11] | 990 (409) [11] |
| $AUC_{inf}$ (mg●day/L) § | 2771 (1549) [10] | 2143 (1316) [10] |
| $C_{28}$ (mg/L) ‖,¶ | 30.4 (11.9) [83] | 24.6 (9.65) [84] |
| Half-life (day) | 32.4 (9.48) [10] | 27.0 (7.57) [10] |

*1200 mg of REGEN-COV contains 600 mg casirivimab and 600 mg of imdevimab.
†Mean (SD) [N]. PK denotes pharmacokinetics, SC subcutaneous, and SD standard deviation.
‡Median (range) [N].
§Value reported for participants with %$AUC_{inf}$ extrapolated <20%.
‖Observed concentration 28 days after dosing, i.e., on Day 29, as defined in the protocol.
¶Value represents participants in the sentinel and safety groups.
